# The genomic epidemiology of SARS-CoV-2 variants of concern in Kenya

**DOI:** 10.1101/2022.10.26.22281446

**Authors:** George Githinji, Arnold W. Lambisia, Ifeanyi Omah, Aine O’Toole, Khadija Said Mohamed, Zaydah R. de Laurent, Timothy O. Makori, Mike Mwanga, Maureen W. Mburu, John M. Morobe, Edidah M. Ong’era, Leonard Ndwiga, Kimita Gathii, Kelvin Thiongo, Don Williams O. Omuoyo, Edith Chepkorir, Jennifer Musyoki, Leonard Kingwara, Damaris Matoke, Samuel O. Oyola, Clayton Onyango, John Waitumbi, Wallace Bulimo, Samoel Khamadi, John N. O. Kiiru, Samson Kinyanjui, Matthew Cotten, Benjamin Tsofa, Isabella Ochola-Oyier, Andrew Rambaut, D. James Nokes, Philip Bejon, Charles Agoti

## Abstract

The emergence and establishment of SARS-CoV-2 variants of concern presented a major global public health crisis across the world. There were six waves of SARS-CoV-2 cases in Kenya that corresponded with the introduction and eventual dominance of the major SARS-COV-2 variants of concern, excepting the first 2 waves that were both wild-type virus. We estimate that more than 1000 SARS-CoV-2 introductions occurred in the two-year epidemic period (March 2020 – September 2022) and a total of 930 introductions were associated with variants of concern namely Beta (n=78), Alpha(n=108), Delta(n=239) and Omicron (n=505). A total of 29 introductions were associated with A.23.1 variant that circulated in high frequencies in Uganda and Rwanda. The actual number of introductions is likely to be higher than these conservative estimates due to limited genomic sequencing. Our data suggested that cryptic transmission was usually underway prior to the first real-time identification of a new variant, and that multiple introductions were responsible. Following emergence of each VOC and subsequent introduction, transmission patterns were associated with hotspots of transmission in Coast, Nairobi and Western Kenya and follows established land and air transport corridors. Understanding the introduction and dispersal of major circulating variants and identifying the sources of new introductions is important to inform public health control strategies within Kenya and the larger East-African region. Border control and case finding reactive to new variants is unlikely to be a successful control strategy.

## Main Text

The emergence and evolution of SARS-CoV-2 in the last 2 years has resulted in extensive global lineage diversity (O’Toole et al., 2021; Rambaut et al., 2020) (n>1,700). This diversity is substantially accounted for by variants with capacity for increased transmission and immune escape, designated as the variants of concern (VOC) by the World Health Organization (WHO) (Konings, Perkins et al. 2021) and comprise of Alpha (du Plessis, McCrone et al. 2021), Beta (Tegally, Wilkinson et al. 2021), Gamma (Faria, Mellan et al. 2021), Delta (Mlcochova, Kemp et al. 2021) and Omicron (Viana, Moyo et al. 2022). VOCs are associated with increased transmissibility (Campbell, Archer et al. 2021, Davies, Abbott et al. 2021, Liu, Liu et al. 2021) and varying degrees of immune escape (Meng, Kemp et al. 2021, Mlcochova, Kemp et al. 2021, Tegally, Wilkinson et al. 2021, Wall, Wu et al. 2021, Hu, Peng et al. 2022). In Kenya, the first confirmed case of SARS-CoV-2 infection was reported on 12^th^ March 2020, and we reported the genomic evidence of multiple early introductions contributing to the first wave of wild-type infections (Githinji, de Laurent et al. 2021). Further introductions occurred in the background of ongoing local transmission of SARS-CoV-2 and spread of the wild-type virus to new populations within Kenya during the second wave (Agoti, Ochola-Oyier et al. 2022).

Kenya experienced a third wave of infections between January and February 2021, a period that coincided with emergence and establishment of Beta and Alpha VOCs in multiple parts of the world and in Kenya. Between June and September 2021, an additional fourth wave of infections was observed that coincided with the increased frequency of the Delta variant and replacement of Alpha and Beta variants. A fifth wave started in late November 2021 driven by the introduction of the Omicron variant. The introduction of BA.2, BA.4 and BA.5 led to a further increase in infections in Kenya and comprised the sixth wave of infection.

To examine the introduction, dispersal, and genomic epidemiology of SARS-CoV-2 variants of concern in Kenya, we analysed sequenced consensus genomes obtained between March 2020 and September 2022. We used these data in conjunction with 54,980 global sequences sampled from over 150 countries and that were most similar to the Kenyan sequences. We used these data to infer the geographical origin of VOC introduction events in Kenya and to estimate the number and arrival dates and subsequent dispersal of multiple lineages between counties within Kenya.

## Materials and Methods

### Ethical Statement

All the analysed samples were collected under the Ministry of Health (MoH) protocols as part of the national response to the COVID-19 pandemic. The whole genome sequencing study protocol was reviewed and approved by the Scientific and Ethics Review Committee (SERU), Kenya Medical Research Institute (KEMRI), Nairobi, Kenya (SERU #4035). Individual patient consent was not required by the committee for the use of these samples for genomic surveillance to inform public health response.

### Study Population

We analysed samples collected across Kenya by the RRT teams from the MoH or those that were collected by private hospitals as part of the government testing and diagnostic strategy. For the samples that were tested at KWTRP, NP/OP swabs were delivered in cool boxes within 48 houses of collection. Samples from outside of the coastal region were only provided for the purpose of whole genome sequencing and were selected based on PCR cycle threshold score (<34). All samples were collected from individuals based on the MoH testing eligibility criteria and this has varied across the two years of the epidemic. An additional set of sequences(n=) were downloaded from GISAID.

### SARS-CoV-2 diagnostics at KWTRP

Viral RNA was extracted from the NP/OP samples and presence of SARS-CoV-2 RNA was confirmed using multiple commercial kits. Cycle threshold cut-offs were as described in the respective kit.

### ONT SARS-CoV-2 library preparation and sequencing

Samples were considered for sequencing if they had PCR cycle threshold value less than 30. Viral RNA from positive samples was extracted using QIAamp Viral RNA Mini kit following the manufacturer’s instructions and reverse transcribed using LunaScript® or Superscript IV RT SuperMix Kit. The cDNA was amplified using Q5® Hot Start High-Fidelity 2x Mastermix with ARTIC nCoV-2019 version 3 primers. The PCR products were run on an agarose gel and successive SARS-CoV-2 to select successful amplification. Successful amplifications were purified using Agencourt AMPure XP beads for library preparation. Sequence libraries were constructed using Oxford Nanopore Technology (ONT) ligation sequencing kit and the ONT Native Barcoding Expansion pack as described in the ARTIC protocol. In every MinION run multiple samples were multiplexed in addition to a no-template control to detect cross contamination. Fast5 files were then base-called using the high accuracy mode available in ONT Guppy base-calling toolkit. The consensus sequences obtained using an ARTIC bioinformatics pipeline. A minimum threshold of x20 reads was required to call a base in the consensus genome else it was masked with an N.

Between December 2020 and May 2022, there were more than three hundred and thirty thousand confirmed cases of SARS-CoV-2 in Kenya reported from across 220 testing centres. A total of 7,266 samples were sequenced at KWTRP and additional samples were sequenced at the ILRI (n=2,598), CDC-Kisumu (n=1,123) and the rest from other KEMRI centres (n=216) making a total of total 11,203 sequences, and which represented 3.4 % of the confirmed cases.

### Lineage and clade assignment

PANGO lineages were assigned to the consensus genomes using the Phylogenetic Assignment of named Global Lineages (PANGOLIN toolkit version 4.1.2. The VOC assignment were based on Scorpio (version 0.3.17) with Constellations v0.1.10. Sequences with low default coverage and that could not be assigned a lineage were excluded from analysis. Clade assignment was done using the command line build of the NextClade tool (version 1.11.0).

### Global SARS-CoV-2 datasets

A total of 8,585,947 sequences and metadata were downloaded from the GISAID (Shu and McCauley 2017) repository as of 19^th^ September 2022. The files were pre-processed by reformatting the sequence headers and the metadata fields. The data was processed as follows. An established SARS-CoV-2 Augur pipeline was used to process the samples based on sampling from the Kenyan and global sequences as summarised in supplementary table 2.

For each of the Kenyan VOC sequences, the corresponding contextual metadata was selected. The Delta lineage comprised of 3 major sub-lineages and each of these were analyzed separately. The A.23.1 and the VOI were also analysed separately. Each group of sequences were aligned using NextAlign version 2.4.0 and the 100bp 5’ and 50bp 3’ ends were masked from the alignment.

We created multiple datasets for analysis depending on the scope (supplementary table 1). In the first dataset, we selected all Kenyan sequences that passed the criteria and was available publicly and in the subsequent datasets were selected depending on the variant type and lineage of interest prevalent in Kenya.

**Table 1:** A summary table showing time of first global detection and VOC detection in Kenya. A detailed summary of first detection of cases in each county is provided in supplementally table1. *Retrospective sequencing of cases from Aga Khan Hospital. ^$^BA.2 could have been circulating in low frequency for several weeks before it became dominant in Europe and South Africa.

| Variant of concern | Date of first global report | Date of first detection in Kenya | Clinical characteristics of initial report | No. of days since global report | No. of introductions |
| --- | --- | --- | --- | --- | --- |
| **Alpha** | 20<sup>th</sup> March 2020 | 29<sup>th</sup> December 2020 | Symptomatic (Fever, headache) | 185 days | 108 |
| **Beta** | 26<sup>th</sup> June 2020 | 15<sup>th</sup> December 2020 | Asymptomatic | 172 days | 78 |
| **Delta** | 3<sup>rd</sup> November 2020 | 7<sup>th</sup> January 2021* | Asymptomatic | 65 days | 239 |
| **Omicron-BA.1** | 17<sup>th</sup> October 2021 | 14<sup>th</sup> November 2021 | Symptomatic | 41 days | 308 |
| **Omicron BA.2** | 22<sup>nd</sup> March 2022<sup>§</sup> | 14<sup>th</sup> December 2021 | Unknown | Sporadic reports<sup>§</sup> | 69 |
| **BA.4** | 14<sup>th</sup> December 2021 | 15<sup>th</sup> April 2022 | Asymptomatic | 122 days | 64 |
| **BA.5** | 15<sup>th</sup> November 2021 | 7<sup>th</sup> April 2022 | Asymptomatic | 143 days | 64 |

### Phylogenetic analysis

Phylogenetic analysis of Kenyan and global sequences was carried using an augur pipeline version 18 using a similar setup described previously by the NextStrain nCoV (https://github.com/nextstrain/ncov). A global context for each variant was specified in within the parameters file. After filtering and quality control a maximum likelihood tree was estimated using IQTree (Minh, Schmidt et al. 2020) version 2.1.4 using a GTR model. The estimated phylogeny was rooted using the reference sequence Wuhan/Hu-1 (GenBank accession MN908947, RefSeq NC_045512.2.3). The maximum likelihood tree was used to estimate a time-scaled phylogenetic tree using TreeTime (Sagulenko, Puller et al. 2018) under a coalescent model. The resulting respective tree was visualized using ggtree package version 3.3.2. in R version 4.4.1.

### Estimating the number of introductions of VOCs into Kenya

We estimated the number of introductions of each of the variants of concern and variants of interest using several methods. We identified Kenyan sequences that did not fall into a Kenyan monophyletic or transmission group as a Kenyan singleton. We inferred that each such singleton comprised of an introduction. In the second approach, the global ML tree topology was used to estimate the number of viral transmission events within the country (Wilkinson, Giovanetti et al. 2021) assuming a contestant evolutionary rate of 8.4 x10-4 nucleotide substitutions per site per year. A mugration model was fitted to the time-scaled phylogenetic tree and the location trait was inferred from the internal nodes. The number of transitions within Kenyan counties were inferred based on the location and inferred date at internal nodes.

## Results

SARS-CoV-2 infections in Kenya were characterised by waves of infections (Figure 1) despite the implementation of control measures such as compulsory masks and restriction of movement and social gatherings among other measures (supplementary figure 1). Each VOC was introduced and sequentially replaced by new variants and in some instances, we observed sequential replacement of sub-lineages (Figure 2) across geographical regions. To provide a global context for variants of concern in Kenya, we carried out phylogenetic and phylodynamic analysis of sequences collected from Kenya with 54,980 global sequences. Kenyan SARS-CoV-2 variants of concern were dispersed throughout the global phylogeny and represented the major diversity that was observed across the globe (Figure 3). The gamma variant was the only major variant that was not reported in Kenya and was also not reported in Africa. Overall diversity comprised of 142 distinct PANGO lineages from sequences collected between March 2020 and September 2022.

**Figure.1:**
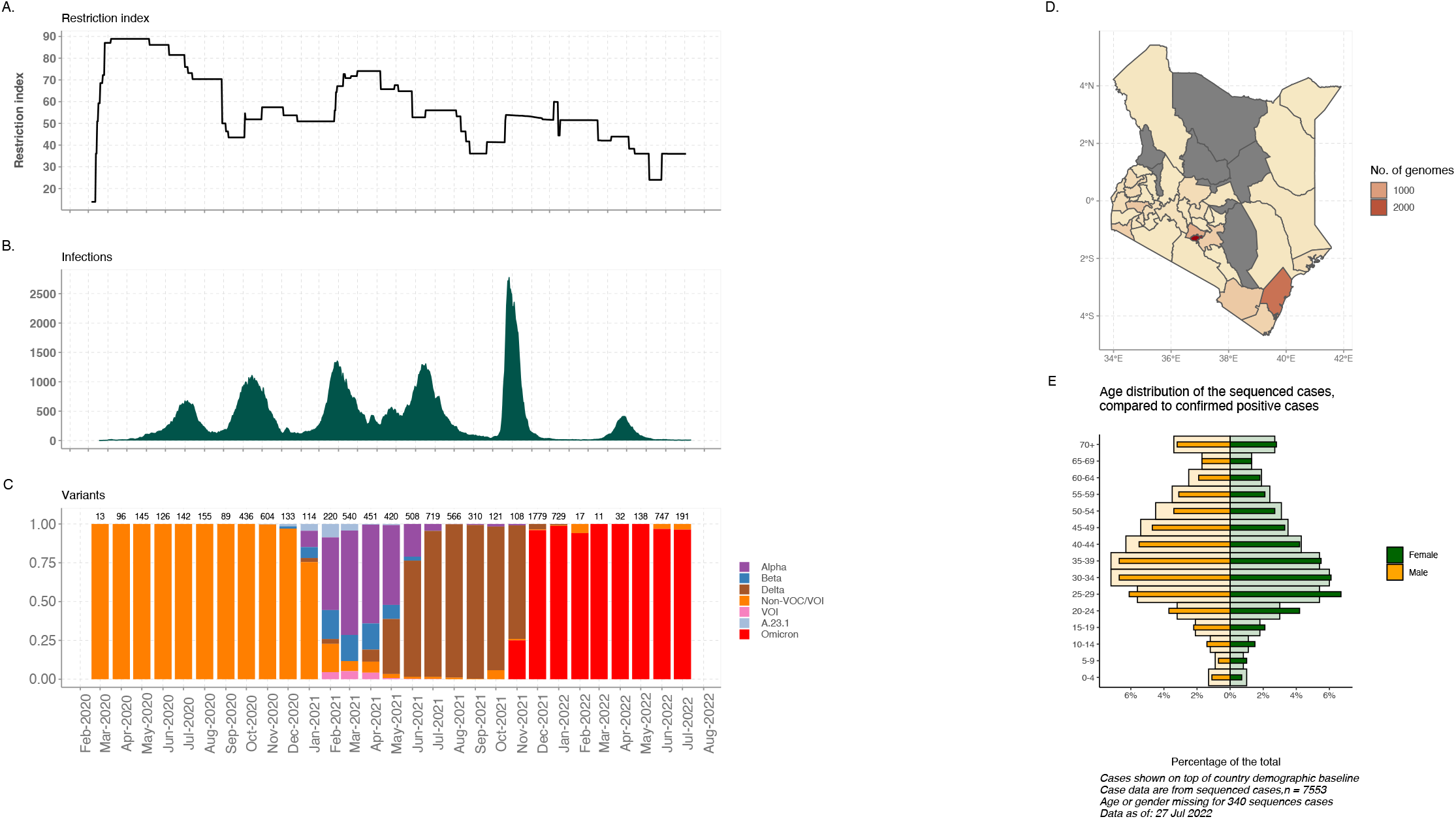
Epidemiological and lineage dynamics of SARS-CoV-2 in Kenya. **A**. A plot showing the Oxford composite restriction index for Kenya between March 2020 and September 2022. **B**. A plot showing the daily number of SARS-CoV-2 cases over the cause of the epidemic in Kenya characteristics by waves of infections between March 2020 and September 2022. **C**. The lower plot shows the major variants associated with each of the wave. Colour denotes each of the variants including the variants of concern and variants of interest circulating at the time. **D**. A map of Kenya showing the counties and the distribution of sequenced genomes per county. The major roads and points of entry are indicated. E. An age distribution plot of the percentage of sequenced cases compared to the percentage of the confirmed SARS-CoV-2 positive cases.

**Figure 2:**
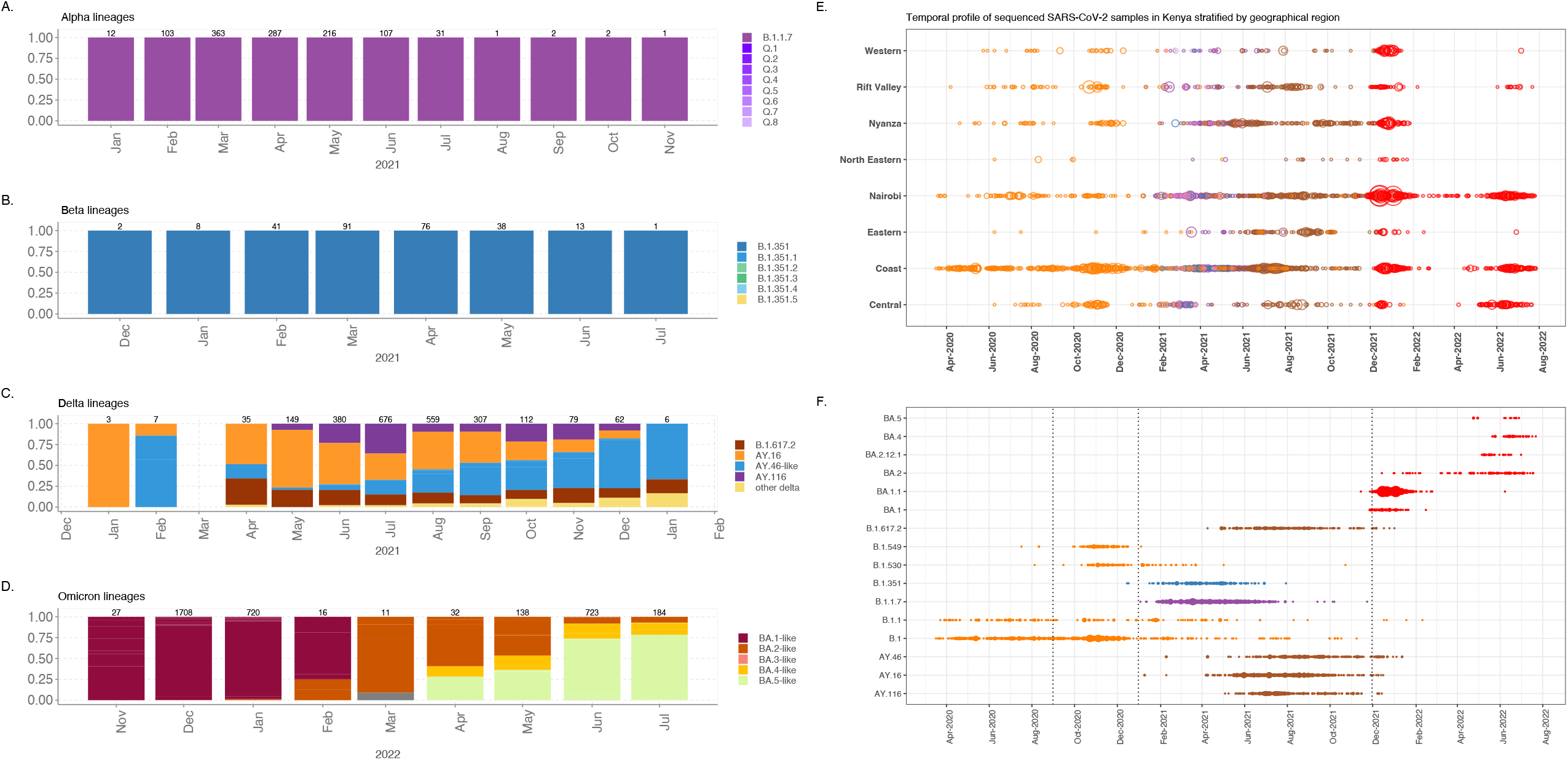
A bar plot showing a summary of the temporal profile for each of the VOC and the respective sub-lineages in Kenya and the normalised temporal profile of the circulating lineages for each for each of the VOCs across the globe that were circulating at the time. **B**. A bubble plot summary of all the major circulating SARS-CoV-2 lineages and sub-lineages in Kenya, between December 2020 and September 2022.

**Figure 3:**
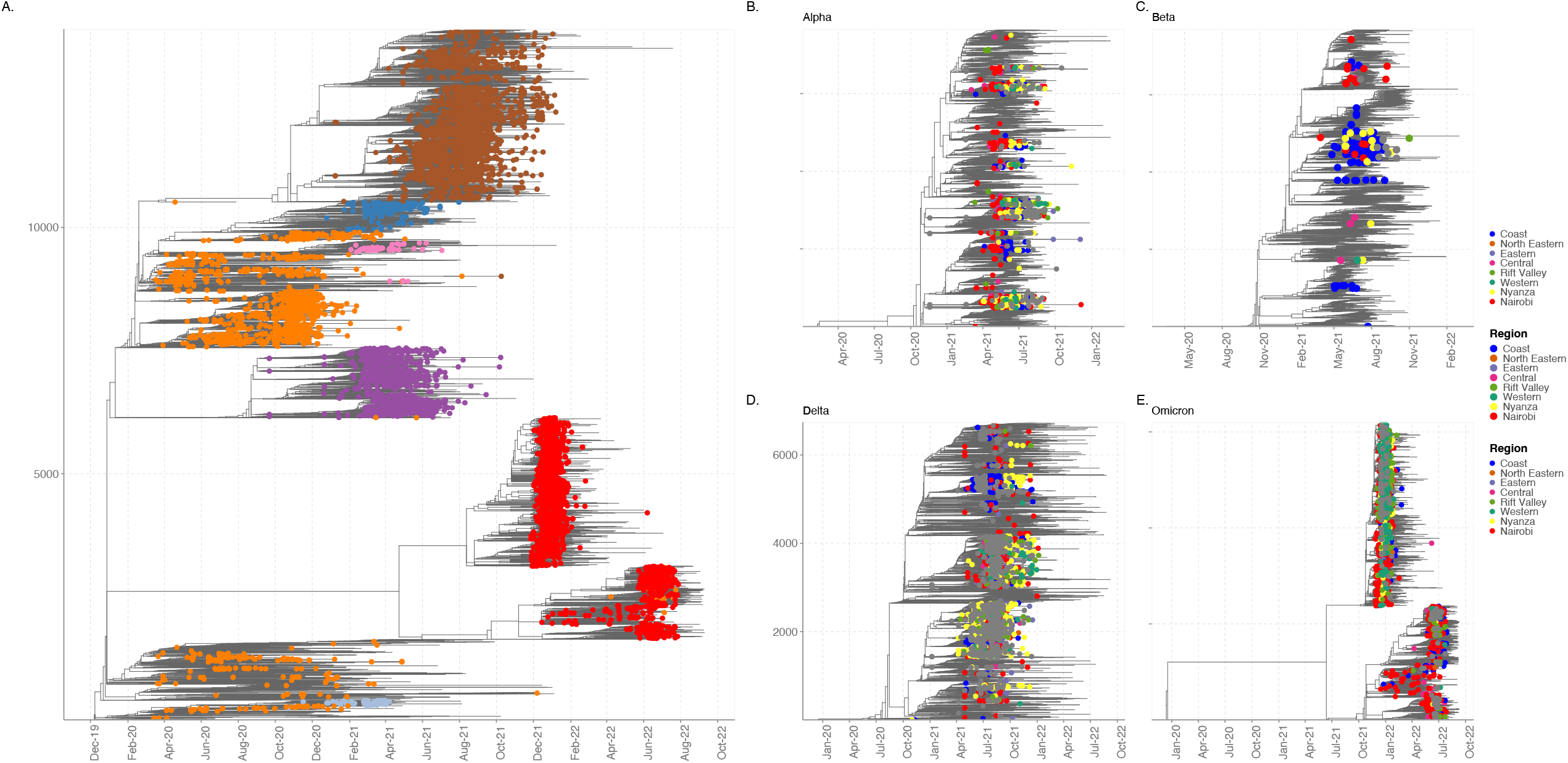
Time resolved maximum likelihood phylogeny of Kenyan SARS-CoV-2 genomes (n=8,438) against a background global SARS-CoV-2 genomes (n=5,592) collected from 184 countries. The clustering of variants of concern, Alpha, Beta, Delta, and Omicron lineages circulating in Kenya are shown in respectively. The A.23.1 lineage circulating in Uganda and Kenya is denoted in xxx. All other lineages are shown in grey.

The Beta variant was the first VOC to be detected in Kenya from samples collected on the 15^th^ and 17^th^ December 2020 from two asymptomatic visitors who had recently travelled from South Africa (Table1). Additional cases of the Beta variant were later detected from individuals who presented at the points of entry (PoE) between Kenya and Tanzania. We identified twenty-four cases of potential direct imports of the Beta variant from individuals with a history of travel from Tanzania. Additional cases were identified from individuals with a history of travel to Tanzania, India, Uganda, and Congo. We analysed a total of 199 sequences from Kenya that were detected between December 2020 and July 2021 and estimated 78 introductions of the Beta variant into Kenya. A few introductions seeded community transmission at the Kenyan coast including outbreaks at the local prisons in Taita-Taveta, Mombasa and Migori counties. The Beta variant was more dominant at the coastal region and only a few cases were detected in other regions (figure 3). This could be attributed to paucity of data or low level of genomic surveillance in other regions relative to the coastal region.

The Alpha variant was reported from routine genomic surveillance of samples collected between 29^th^ December 2020 and 14^th^ January 2021 (Table 1). The first Alpha variant was from a male individual (less than 35 years old) who presented to a Mombasa clinic with symptoms of fever and headache and without a history of recent international travel or known contact with an infected individual, suggesting previous cryptic transmission of this variant within the population prior to isolation. We identified a total of 1,101 Alpha variant sequences and based on these we estimate at least 102 introductions into the country. Three introductions were associated with large community outbreaks (n>100) cases and 13 further introductions were associated with transmission clusters of 10 or more cases. Alpha cases in Kenya represented most of the African and global diversity. For example, we noted a large transmission cluster of Western African origin that was introduced into Nairobi.

The delta variant outbreak was observed in real time among samples that were tested in April 2021 in Kisumu and Nairobi from individuals with a history of travel to/from India. Further retrospective sequencing of stored samples identified cases of the Delta variant as early as 9^th^ January 2021 in Kilifi on the Kenyan Coast (n=3), and an additional 7 cases were diagnosed at the Nairobi Aga-Khan hospital on February 9^th^, 2021. All the 10 cases came from individuals who had no history of recent international travel, suggesting earlier cryptic transmission. At the Coast the first reported case was in Mombasa from an individual with history of recent travel to India. In addition, on 29^th^ April 2021 we reported additional cases through airport surveillance of cases from individuals with history of travel from India. We estimated at least 380 introductions from 2,527 fully sequenced Delta variant genomes. The Delta variant replaced the Alpha variant in Kenya by June 2021. The Delta variant was shown to have been more transmissible than the Alpha variant. There were four major Delta sub lineages associated with multiple infections in Kenya i.e., AY.116, AY.16, AY.46 and B.1.617.2 and at least 28 further additional lineages that did not lead to significant further transmission in Kenya.

The first genomic detection of Omicron cases was from samples collected in Nairobi on 14^th^ November 2021 (Table 1). This was identified retrospectively by sequencing of samples stored by the National Public Health Laboratory (NPHL) and was from an individual without prior history of travel, providing evidence that the Omicron variant was circulating in Kenya prior to the initial real time detection. The number of cases rose from 20 to a peak of 3000 confirmed cases per day by January 2022. This was followed by a sudden decline in the number of cases by end of February 2022, and a change of policy on testing and the introduction of antigen tests which further impacted the number of samples available for sequencing.

The BA.1 co-circulated with BA.1.1. variants which were the most dominant of the two lineages driving the 5^th^ wave. Despite an increase in the number of BA.2 variants globally and a BA.2 driven wave of cases in South Africa, there was no remarkable increase in the number of cases and BA.2 variants did not cause a major wave in Kenya. Between May and July 2022, a sixth wave of infections associated with increased circulation BA.2.12.1, BA.4 and BA.5 was observed. The emergence of BA.4 first detected on 14^th^ December 2021 and BA.5 first detected on 15^th^ November 2021, in South Africa.

### Temporal replacement of variants of concern

Our previous studies showed that the first and second waves were characterised by introductions of the virus from the global community (Githinji, de Laurent et al. 2021) followed by establishment and sustained local transmission of the wildtype strains (Agoti, Ochola-Oyier et al. 2021) which occurred within different demographic populations and social-economic groups (Brand, Ojal et al. 2021). Here we show that subsequent waves coincided with multiple introductions of two variants of concern (i.e., Alpha and Beta) and a variant of interest between January and May 2021 while the fourth wave coincided with the establishment of the Delta variant between April and September 2021 (figure 1). The fifth wave coincided with multiple introductions and rapid transmission of the Omicron variant and the sixth wave corresponded with introductions of the BA.4 and BA.5 omicron lineages (figure 1 and figure 3).

Upon introduction, each VOC established rapid local transmission despite implementation of control measures such as wearing of masks and control of movement past evening hours, and there was evidence that local transmission was already established prior to the point of first identification. Both Beta and Alpha variants co-circulated in Kenya for a couple of weeks following introduction of the Alpha variant (Figure 1 and figure 3). To gain insight on local patterns of spread, we generated both maximum likelihood and time-resolved phylogenetic trees from sequences collected between March 2020 and September 2022, our study focuses more on the sequences collected between late 2020 to September 2022.

Our data suggests that each of the major VOC showed distinct pattern of introduction and dispersal within the country. The Beta variant was prevalent along the coastal counties (figure 3). A total of 18 of these cases were sequenced from samples collected at the entry points in Kwale and Taita-Taveta counties from truck drivers between January and March 2021 and 23 cases were recorded among inmates at two prison facilities in Mombasa and Taita-Taveta. And additional 12 cases were identified at a prison facility in Western Kenya. Overall, our data suggests that the coastal region was an important region of entry and dispersal point for Beta variants to the rest of the country.

Nairobi region and its environs may have been a major dispersal point for the Alpha variant to the rest of the country (supplementary figure 2). Other significant regions of dispersal were the coastal and western Kenya. The Alpha variant showed geographical clustering and was more dispersed relative to the Beta variant. Nairobi was a major dispersal region (supplementary figure 2) to coast, central and western Kenya regions. On the other hand, the Delta variant showed distinct geographical and variant clustering (figure 3). Each of these lineages were dispersed to other regions with the Nyanza region acting as a major source of within country transmission (supplementary figure 2). Upon introduction in late 2021, the Omicron BA.1 and BA.1.1 were the most dominant variants and appear to have been largely dispersed from Nairobi. The two variants were associated with widespread infections in Nairobi and its environs. Nairobi was a major source of the BA.1.1 for other regions in the country. Kenya did not see large widespread or domination of the BA.2 variant, nevertheless, an increase in the number of BA.2, BA.4 and BA.5 was associated with the 6^th^ wave of infections. Nairobi region was a source of the BA.5 variants.

## Discussion

The evolution of SARS-CoV-2 has been characterised by emergence of multiple lineages with capacity for increased transmission and immune escape. These variants of concern (VOC) spread swiftly across the globe replacing the previous wild-type strains. We document here the sequential introduction of these VOC lineages in Kenya, except for the Gamma variant that was widely reported in South and North America and some regions in Europe. Seroprevalence data integrated with transmission modelling (Brand, Ojal et al. 2021) indicated that a high proportion of the Kenya population had been exposed to the wild-type SARS-CoV-2 virus by the end of the second wave in December 2020 (Ojal, Brand et al. 2020, Adetifa, Uyoga et al. 2021, Etyang, Lucinde et al. 2021, Uyoga, Adetifa et al. 2021, Uyoga, Adetifa et al. 2021). The third wave coincided with the introduction of VOCs which suggests a role for the Beta and Alpha variants in the third wave. The fourth wave followed the introduction and spread of the Delta VOC in May 2021. The fifth wave followed the introduction of the highly transmissible Omicron variant and led to the largest spike in infected cases. A sixth wave was driven by fitter subvariants of the Omicron wave namely BA.2, BA.4 and BA.5 variants that were introduced in March 2022. It is difficult to ascertain the total number of omicron BA.4 and BA.5 infections given that the testing capacity in Kenya dropped significantly since March 2022. Natural immunity from previous infections and vaccination could have played a significant role in the number of actual cases that were observed. The variants of concern were a major driver of the surges in cases.

We find that cryptic transmission of VOCS occurred prior to initial genomic detection which underscores the limited genomic surveillance of SARS-CoV-2 in Kenya against a background of reactive public health interventions such as restriction of movement, ban on public gatherings and mask mandate. By the time these efforts were made, it was too late, and the virus was already well established within the respective community. We find that the time to first genomic detection of later variants such as omicron was lower relative to Beta and Alpha variant. The latter were introduced at a time of limited public health measures.

Surveillance effort should focus in identifying cases of cryptic transmission among venerable groups e.g., immune-compromised groups. Recent convergent evolution of multiple Omicron variants should be investigated among cases in Kenya. In addition, understanding of the factors that contributed to the growth or emergence of transmission clusters in specific regions or among specific demographic groups would be essential to inform future control measures. Continued surveillance of SARS-CoV-2 variants in Kenya is recognized by the Ministry of Health, Government of Kenya, as important in forecasting the health resource needs and for monitoring of emerging local diversity of VOC with further potential for increased transmissibility or immune escape, particularly against the background of a large population characterized by partial immunity.

## Data Availability

All data produced are available online at https://github.com/george-githinji/VOC_manuscript

## Acknowledgements

We thank (a) the members of the six Coastal counties of Kenya RRTs for collecting the samples analysed here; (b) the members of the COVID-19 KWTRP Testing Team who tirelessly analysed the samples received at KWTRP to identify positives (see full list of members below); (c) the KWTRP data entry team, (d) Laboratories that have shared sequence data on GISAID that we included as comparison data in our analysis (see list in appendix); (e) the KRISP team in South Africa for sharing the scripts we used in the import/export analysis and AFRICA-CDC for facilitating Africa genomics training. This paper is published with permission of the Director of KEMRI.

## Members of COVID-19 Testing Team at KWTRP

Agnes Mutiso, Alfred Mwanzu, Angela Karani, Bonface M. Gichuki, Boniface Karia, Brian Bartilol, Brian Tawa, Calleb Odundo, Caroline Ngetsa, Clement Lewa, Daisy Mugo, David Amadi, David Ireri, Debra Riako, Domtila Kimani, Edwin Machanja, Elijah Gicheru, Elisha Omer, Faith Gambo, Horace Gumba, Isaac Musungu, James Chemweno, Janet Thoya, Jedida Mwacharo, John Gitonga, Johnstone Makale, Justine Getonto, Kelly Ominde, Kelvias Keter, Lydia Nyamako, Margaret Nunah, Martin Mutunga, Metrine Tendwa, Moses Mosobo, Nelson Ouma, Nicole Achieng, Patience Kiyuka, Perpetual Wanjiku, Peter Mwaura, Rita Warui, Robinson Cheruiyot, Salim Mwarumba, Shaban Mwangi, Shadrack Mutua, Sharon Owuor, Susan Njuguna, Victor Osoti, Wesley Cheruiyot, Wilfred Nyamu, Wilson Gumbi and Yiakon Sein.

## Acknowledgments

