## Supplementary figures and images for "The genomic epidemiology of SARS-CoV-2 variants of concern in Kenya"

### supplementary figure 1

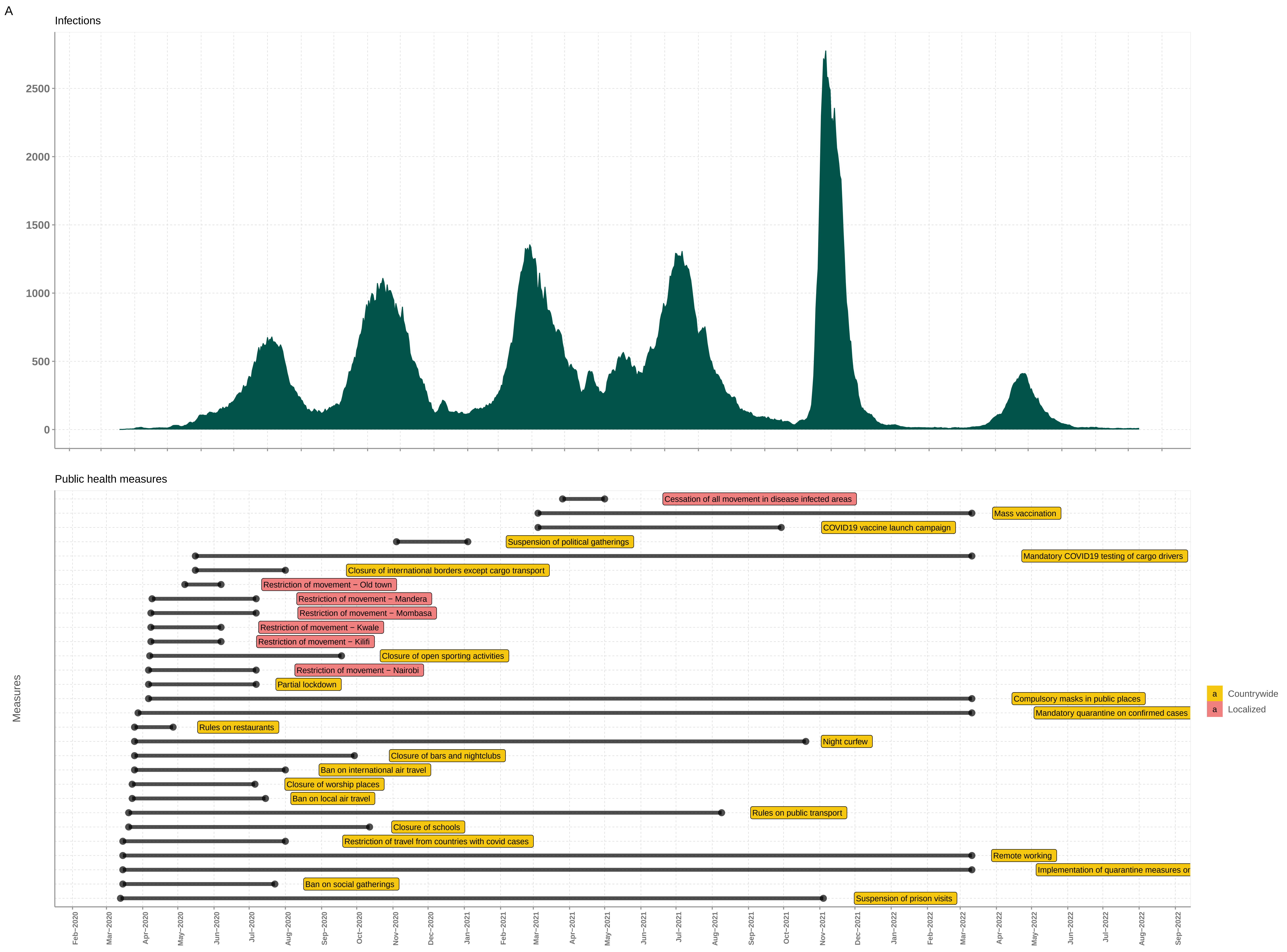

### supplementary figure 2

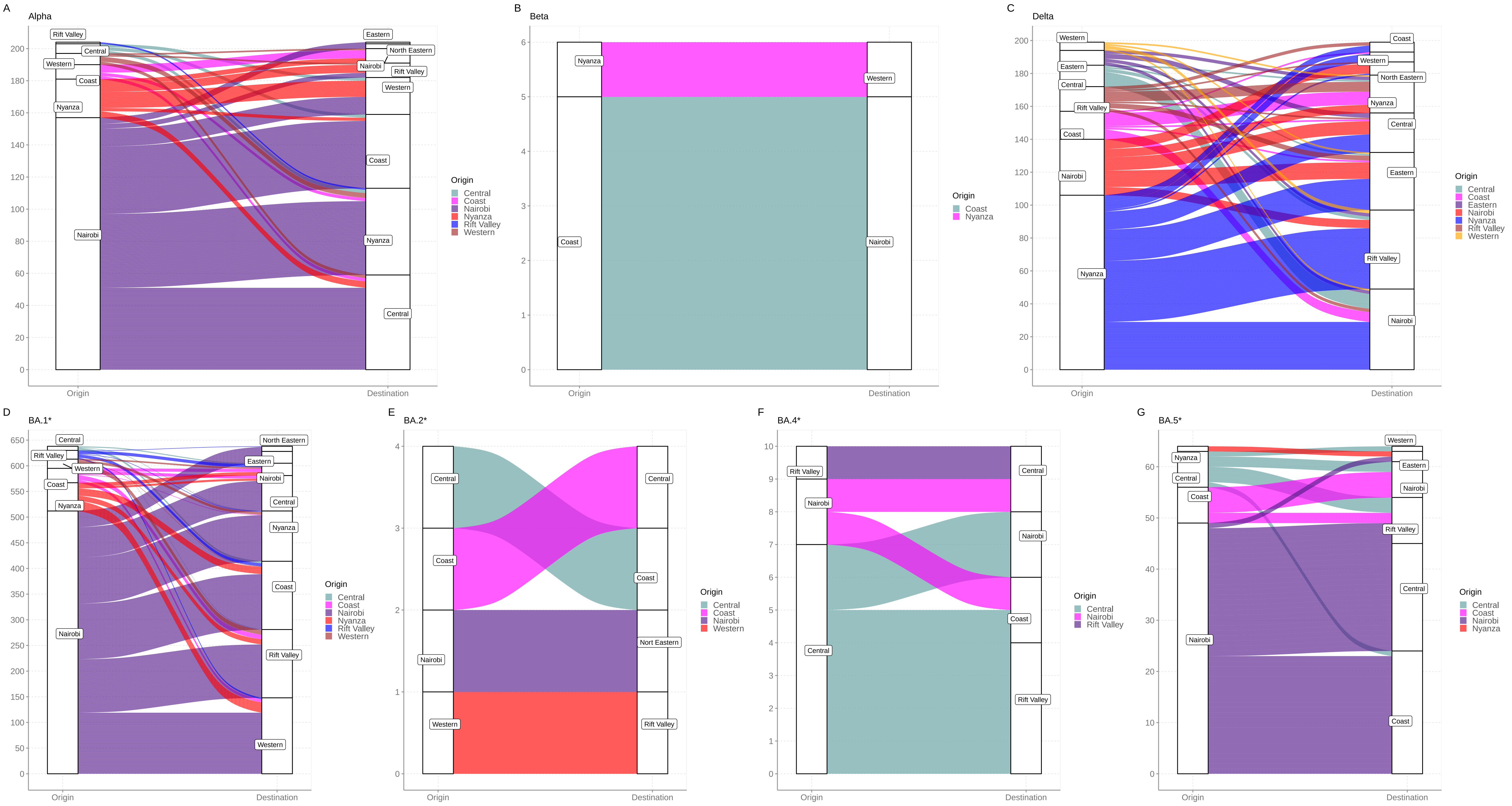

### supplementary figure 3

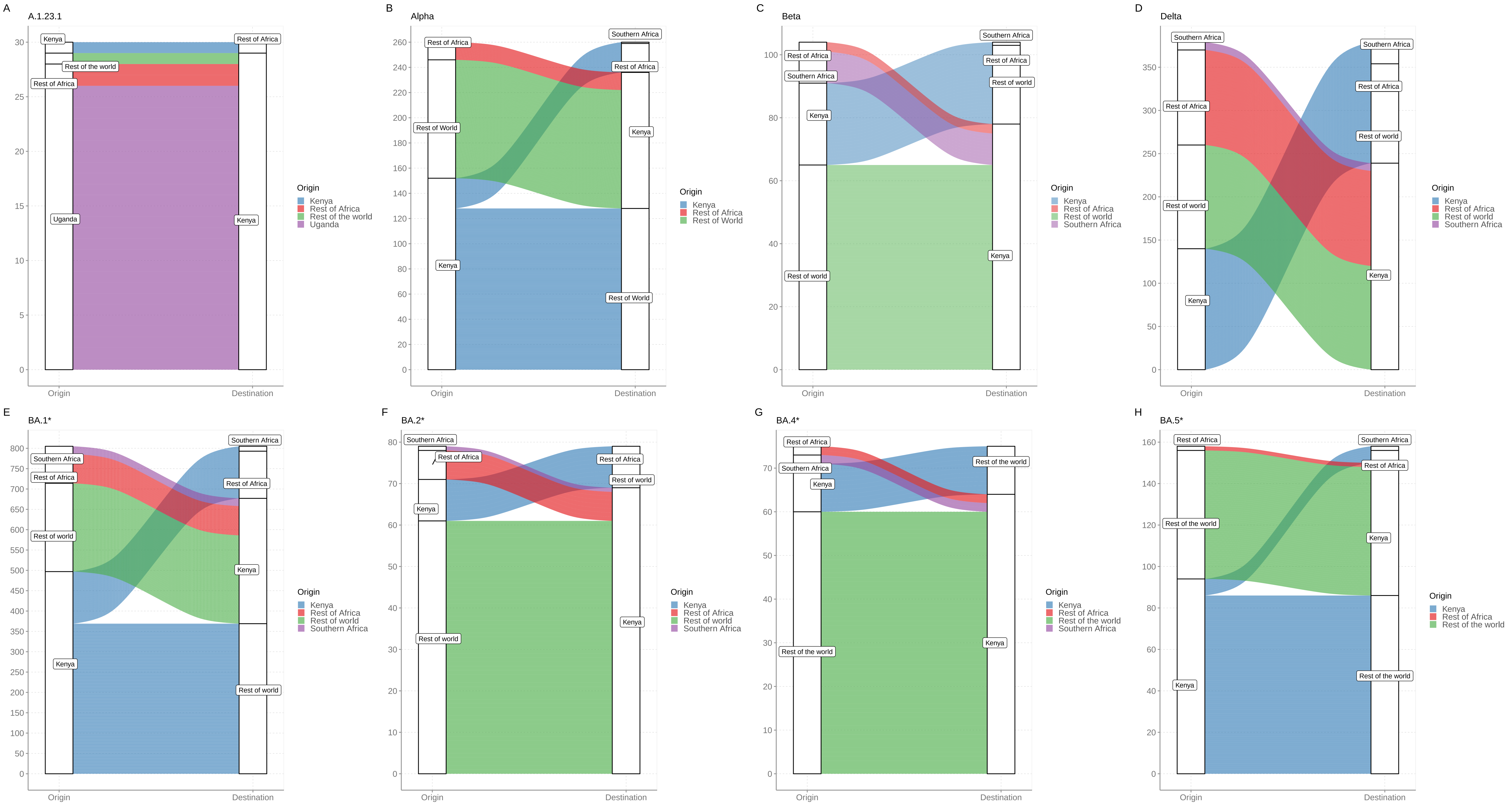
