## supplementary table 1 for "The genomic epidemiology of SARS-CoV-2 variants of concern in Kenya"

| Variant | Lineage | No. of Kenyan sequences | No. of sequences from the global context | No. of countries in the global dataset |
| --- | --- | --- | --- | --- |
| Alpha | B.1.1.7 | 1,039 | 8,539 | 96 |
| Beta |  | 199 | 7,503 | 83 |
| Delta | Delta lineages | 2,301 | 4,438 | 168 |
|  | AY.16 | 1,049 | 2,802 | 43 |
|  | AY.46 | 582 | 5,557 | 76 |
|  | AY.116 | 348 | 2,826 | 62 |
| Omicron | Omicron lineages | 2,931 | 3,267 | 167 |
|  | BA.1 | 2,298 | 5,131 | 156 |
|  | BA.2 | 134 | 4,679 | 138 |
|  | BA.4 | 146 | 5,142 | 100 |
|  | BA.5 | 425 | 5,096 | 118 |
| All Kenyan lineages | All lineages | 8,438 | 5,592 | 184 |

**Supplementary table 1:** A summary of the Kenyan and global sequences used for phylogenetic analysis and to estimate the number of introductions based on the global context.
