## supplementary table 2 for "The genomic epidemiology of SARS-CoV-2 variants of concern in Kenya"

| **lineage** | **county** | **date_collected_epi** | **most_common_countries** | **Earliest date of detection** | **days_since_global_report** |
| --- | --- | --- | --- | --- | --- |
| AY.1 | Kilifi | 2021-07-06 | United States of America 71.0%, Philippines 8.0%, India 4.0%, Japan 2.0%, United Kingdom 2.0% | 2021-03-31 | 97 |
| AY.101 | Nairobi | 2021-10-15 | Brazil 60.0%, India 22.0%, Chile 6.0%, Colombia 3.0%, Paraguay 1.0% | 2021-01-11 | 277 |
| AY.103 | Migori | 2021-10-05 | United States of America 97.0%, Canada 1.0%, United Kingdom 0.0%, Mexico 0.0%, Switzerland 0.0% | 2020-10-06 | 364 |
| AY.110 | Mombasa | 2021-06-18 | United States of America 89.0%, Italy 2.0%, United Kingdom 1.0%, Canada 1.0%, Germany 1.0% | 2021-04-14 | 65 |
| AY.116 | Kakamega | 2021-10-07 | Norway 13.0%, Sweden 11.0%, Brazil 10.0%, Kenya 9.0%, Zambia 8.0% | 2021-01-21 | 259 |
| AY.116 | Kilifi | 2021-05-04 | Norway 13.0%, Sweden 11.0%, Brazil 10.0%, Kenya 9.0%, Zambia 8.0% | 2021-01-21 | 103 |
| AY.116 | Kisii | 2021-09-16 | Norway 13.0%, Sweden 11.0%, Brazil 10.0%, Kenya 9.0%, Zambia 8.0% | 2021-01-21 | 238 |
| AY.116 | Kwale | 2021-07-19 | Norway 13.0%, Sweden 11.0%, Brazil 10.0%, Kenya 9.0%, Zambia 8.0% | 2021-01-21 | 179 |
| AY.116 | Lamu | 2021-07-05 | Norway 13.0%, Sweden 11.0%, Brazil 10.0%, Kenya 9.0%, Zambia 8.0% | 2021-01-21 | 165 |
| AY.116 | Machakos | 2021-06-20 | Norway 13.0%, Sweden 11.0%, Brazil 10.0%, Kenya 9.0%, Zambia 8.0% | 2021-01-21 | 150 |
| AY.116 | Migori | 2021-10-06 | Norway 13.0%, Sweden 11.0%, Brazil 10.0%, Kenya 9.0%, Zambia 8.0% | 2021-01-21 | 258 |
| AY.116 | Mombasa | 2021-06-12 | Norway 13.0%, Sweden 11.0%, Brazil 10.0%, Kenya 9.0%, Zambia 8.0% | 2021-01-21 | 142 |
| AY.116 | Nairobi | 2021-05-09 | Norway 13.0%, Sweden 11.0%, Brazil 10.0%, Kenya 9.0%, Zambia 8.0% | 2021-01-21 | 108 |
| AY.116 | Nyamira | 2021-09-13 | Norway 13.0%, Sweden 11.0%, Brazil 10.0%, Kenya 9.0%, Zambia 8.0% | 2021-01-21 | 235 |
| AY.116 | Taita Taveta | 2021-06-10 | Norway 13.0%, Sweden 11.0%, Brazil 10.0%, Kenya 9.0%, Zambia 8.0% | 2021-01-21 | 140 |
| AY.116 | Tana River | 2021-07-06 | Norway 13.0%, Sweden 11.0%, Brazil 10.0%, Kenya 9.0%, Zambia 8.0% | 2021-01-21 | 166 |
| AY.120 | Machakos | 2021-07-29 | United Kingdom 82.0%, India 4.0%, United States of America 3.0%, Germany 2.0%, France 1.0% | 2020-10-26 | 276 |
| AY.120 | Nairobi | 2021-08-22 | United Kingdom 82.0%, India 4.0%, United States of America 3.0%, Germany 2.0%, France 1.0% | 2020-10-26 | 300 |
| AY.122 | Kilifi | 2021-07-07 | Germany 18.0%, United States of America 13.0%, Denmark 10.0%, France 8.0%, Sweden 5.0% | 2020-05-11 | 422 |
| AY.122 | Kisumu | 2021-08-20 | Germany 18.0%, United States of America 13.0%, Denmark 10.0%, France 8.0%, Sweden 5.0% | 2020-05-11 | 466 |
| AY.122 | Kwale | 2021-06-14 | Germany 18.0%, United States of America 13.0%, Denmark 10.0%, France 8.0%, Sweden 5.0% | 2020-05-11 | 399 |
| AY.122 | Lamu | 2021-08-21 | Germany 18.0%, United States of America 13.0%, Denmark 10.0%, France 8.0%, Sweden 5.0% | 2020-05-11 | 467 |
| AY.122 | Machakos | 2021-08-02 | Germany 18.0%, United States of America 13.0%, Denmark 10.0%, France 8.0%, Sweden 5.0% | 2020-05-11 | 448 |
| AY.122 | Migori | 2021-10-04 | Germany 18.0%, United States of America 13.0%, Denmark 10.0%, France 8.0%, Sweden 5.0% | 2020-05-11 | 511 |
| AY.122 | Mombasa | 2021-06-10 | Germany 18.0%, United States of America 13.0%, Denmark 10.0%, France 8.0%, Sweden 5.0% | 2020-05-11 | 395 |
| AY.122 | Nairobi | 2021-08-21 | Germany 18.0%, United States of America 13.0%, Denmark 10.0%, France 8.0%, Sweden 5.0% | 2020-05-11 | 467 |
| AY.122 | Taita Taveta | 2021-08-30 | Germany 18.0%, United States of America 13.0%, Denmark 10.0%, France 8.0%, Sweden 5.0% | 2020-05-11 | 476 |
| AY.126 | Nairobi | 2021-12-07 | Germany 25.0%, Denmark 19.0%, Turkey 15.0%, Sweden 6.0%, Switzerland 5.0% | 2020-10-14 | 419 |
| AY.127 | Nairobi | 2021-09-25 | India 18.0%, United Kingdom 13.0%, Germany 12.0%, Denmark 11.0%, Norway 8.0% | 2021-01-08 | 260 |
| AY.16 | Baringo | 2021-04-16 | India 33.0%, Kenya 27.0%, United States of America 22.0%, Denmark 4.0%, Germany 2.0% | 2021-01-07 | 99 |
| AY.16 | Bomet | 2021-08-02 | India 33.0%, Kenya 27.0%, United States of America 22.0%, Denmark 4.0%, Germany 2.0% | 2021-01-07 | 207 |
| AY.16 | Busia | 2021-05-31 | India 33.0%, Kenya 27.0%, United States of America 22.0%, Denmark 4.0%, Germany 2.0% | 2021-01-07 | 144 |
| AY.16 | Garissa | 2021-08-19 | India 33.0%, Kenya 27.0%, United States of America 22.0%, Denmark 4.0%, Germany 2.0% | 2021-01-07 | 224 |
| AY.16 | Homa Bay | 2021-06-18 | India 33.0%, Kenya 27.0%, United States of America 22.0%, Denmark 4.0%, Germany 2.0% | 2021-01-07 | 162 |
| AY.16 | Kakamega | 2021-09-29 | India 33.0%, Kenya 27.0%, United States of America 22.0%, Denmark 4.0%, Germany 2.0% | 2021-01-07 | 265 |
| AY.16 | Kericho | 2021-06-11 | India 33.0%, Kenya 27.0%, United States of America 22.0%, Denmark 4.0%, Germany 2.0% | 2021-01-07 | 155 |
| AY.16 | Kiambu | 2021-04-18 | India 33.0%, Kenya 27.0%, United States of America 22.0%, Denmark 4.0%, Germany 2.0% | 2021-01-07 | 101 |
| AY.16 | Kilifi | 2021-05-13 | India 33.0%, Kenya 27.0%, United States of America 22.0%, Denmark 4.0%, Germany 2.0% | 2021-01-07 | 126 |
| AY.16 | Kisii | 2021-04-19 | India 33.0%, Kenya 27.0%, United States of America 22.0%, Denmark 4.0%, Germany 2.0% | 2021-01-07 | 102 |
| AY.16 | Kisumu | 2021-04-07 | India 33.0%, Kenya 27.0%, United States of America 22.0%, Denmark 4.0%, Germany 2.0% | 2021-01-07 | 90 |
| AY.16 | Laikipia | 2021-05-26 | India 33.0%, Kenya 27.0%, United States of America 22.0%, Denmark 4.0%, Germany 2.0% | 2021-01-07 | 139 |
| AY.16 | Lamu | 2021-07-18 | India 33.0%, Kenya 27.0%, United States of America 22.0%, Denmark 4.0%, Germany 2.0% | 2021-01-07 | 192 |
| AY.16 | Machakos | 2021-05-28 | India 33.0%, Kenya 27.0%, United States of America 22.0%, Denmark 4.0%, Germany 2.0% | 2021-01-07 | 141 |
| AY.16 | Migori | 2021-06-03 | India 33.0%, Kenya 27.0%, United States of America 22.0%, Denmark 4.0%, Germany 2.0% | 2021-01-07 | 147 |
| AY.16 | Mombasa | 2021-04-25 | India 33.0%, Kenya 27.0%, United States of America 22.0%, Denmark 4.0%, Germany 2.0% | 2021-01-07 | 108 |
| AY.16 | Nairobi | 2021-01-09 | India 33.0%, Kenya 27.0%, United States of America 22.0%, Denmark 4.0%, Germany 2.0% | 2021-01-07 | 2 |
| AY.16 | Nandi | 2021-06-25 | India 33.0%, Kenya 27.0%, United States of America 22.0%, Denmark 4.0%, Germany 2.0% | 2021-01-07 | 169 |
| AY.16 | Nyamira | 2021-09-13 | India 33.0%, Kenya 27.0%, United States of America 22.0%, Denmark 4.0%, Germany 2.0% | 2021-01-07 | 249 |
| AY.16 | Nyeri | 2021-05-25 | India 33.0%, Kenya 27.0%, United States of America 22.0%, Denmark 4.0%, Germany 2.0% | 2021-01-07 | 138 |
| AY.16 | Siaya | 2021-05-17 | India 33.0%, Kenya 27.0%, United States of America 22.0%, Denmark 4.0%, Germany 2.0% | 2021-01-07 | 130 |
| AY.16 | Taita Taveta | 2021-07-21 | India 33.0%, Kenya 27.0%, United States of America 22.0%, Denmark 4.0%, Germany 2.0% | 2021-01-07 | 195 |
| AY.16 | Uasin Gishu | 2021-07-29 | India 33.0%, Kenya 27.0%, United States of America 22.0%, Denmark 4.0%, Germany 2.0% | 2021-01-07 | 203 |
| AY.16 | Vihiga | 2021-06-29 | India 33.0%, Kenya 27.0%, United States of America 22.0%, Denmark 4.0%, Germany 2.0% | 2021-01-07 | 173 |
| AY.36 | Nairobi | 2021-09-25 | United Kingdom 25.0%, Denmark 15.0%, Nigeria 12.0%, Germany 9.0%, United States of America 9.0% | 2020-08-11 | 410 |
| AY.38 | Kilifi | 2021-07-30 | South_Africa 69.0%, Sweden 11.0%, United States of America 4.0%, Germany 3.0%, India 2.0% | 2021-03-27 | 125 |
| AY.39 | Kilifi | 2021-06-17 | United States of America 84.0%, India 4.0%, United Kingdom 4.0%, Australia 2.0%, Germany 1.0% | 2020-10-27 | 233 |
| AY.4 | Nairobi | 2021-08-22 | United Kingdom 84.0%, Denmark 3.0%, Germany 2.0%, Ireland 2.0%, France 1.0% | 2020-05-11 | 468 |
| AY.4.2.2 | Nairobi | 2021-11-16 | United Kingdom 96.0%, Ireland 1.0%, France 0.0%, Sweden 0.0%, Germany 0.0% | 2021-04-12 | 218 |
| AY.44 | Migori | 2021-10-06 | United States of America 98.0%, Canada 1.0%, Germany 0.0%, Turkey 0.0%, United Kingdom 0.0% | 2020-07-14 | 449 |
| AY.44 | Nairobi | 2021-08-28 | United States of America 98.0%, Canada 1.0%, Germany 0.0%, Turkey 0.0%, United Kingdom 0.0% | 2020-07-14 | 410 |
| AY.45 | Machakos | 2021-08-22 | South_Africa 70.0%, United States of America 5.0%, India 3.0%, Sweden 3.0%, United Kingdom 3.0% | 2021-01-09 | 225 |
| AY.46 | Baringo | 2021-09-22 | Denmark 20.0%, United Kingdom 11.0%, Germany 10.0%, Sweden 8.0%, United States of America 7.0% | 2020-06-01 | 478 |
| AY.46 | Bomet | 2021-08-10 | Denmark 20.0%, United Kingdom 11.0%, Germany 10.0%, Sweden 8.0%, United States of America 7.0% | 2020-06-01 | 435 |
| AY.46 | Bungoma | 2021-12-06 | Denmark 20.0%, United Kingdom 11.0%, Germany 10.0%, Sweden 8.0%, United States of America 7.0% | 2020-06-01 | 553 |
| AY.46 | Busia | 2021-06-24 | Denmark 20.0%, United Kingdom 11.0%, Germany 10.0%, Sweden 8.0%, United States of America 7.0% | 2020-06-01 | 388 |
| AY.46 | Garissa | 2021-08-02 | Denmark 20.0%, United Kingdom 11.0%, Germany 10.0%, Sweden 8.0%, United States of America 7.0% | 2020-06-01 | 427 |
| AY.46 | Homa Bay | 2021-12-14 | Denmark 20.0%, United Kingdom 11.0%, Germany 10.0%, Sweden 8.0%, United States of America 7.0% | 2020-06-01 | 561 |
| AY.46 | Kakamega | 2021-09-11 | Denmark 20.0%, United Kingdom 11.0%, Germany 10.0%, Sweden 8.0%, United States of America 7.0% | 2020-06-01 | 467 |
| AY.46 | Kericho | 2021-07-12 | Denmark 20.0%, United Kingdom 11.0%, Germany 10.0%, Sweden 8.0%, United States of America 7.0% | 2020-06-01 | 406 |
| AY.46 | Kiambu | 2021-07-07 | Denmark 20.0%, United Kingdom 11.0%, Germany 10.0%, Sweden 8.0%, United States of America 7.0% | 2020-06-01 | 401 |
| AY.46 | Kilifi | 2021-06-13 | Denmark 20.0%, United Kingdom 11.0%, Germany 10.0%, Sweden 8.0%, United States of America 7.0% | 2020-06-01 | 377 |
| AY.46 | Kisii | 2021-08-26 | Denmark 20.0%, United Kingdom 11.0%, Germany 10.0%, Sweden 8.0%, United States of America 7.0% | 2020-06-01 | 451 |
| AY.46 | Kisumu | 2021-05-12 | Denmark 20.0%, United Kingdom 11.0%, Germany 10.0%, Sweden 8.0%, United States of America 7.0% | 2020-06-01 | 345 |
| AY.46 | Kwale | 2021-08-15 | Denmark 20.0%, United Kingdom 11.0%, Germany 10.0%, Sweden 8.0%, United States of America 7.0% | 2020-06-01 | 440 |
| AY.46 | Laikipia | 2021-06-18 | Denmark 20.0%, United Kingdom 11.0%, Germany 10.0%, Sweden 8.0%, United States of America 7.0% | 2020-06-01 | 382 |
| AY.46 | Lamu | 2021-06-18 | Denmark 20.0%, United Kingdom 11.0%, Germany 10.0%, Sweden 8.0%, United States of America 7.0% | 2020-06-01 | 382 |
| AY.46 | Machakos | 2021-06-05 | Denmark 20.0%, United Kingdom 11.0%, Germany 10.0%, Sweden 8.0%, United States of America 7.0% | 2020-06-01 | 369 |
| AY.46 | Migori | 2021-10-05 | Denmark 20.0%, United Kingdom 11.0%, Germany 10.0%, Sweden 8.0%, United States of America 7.0% | 2020-06-01 | 491 |
| AY.46 | Mombasa | 2021-06-19 | Denmark 20.0%, United Kingdom 11.0%, Germany 10.0%, Sweden 8.0%, United States of America 7.0% | 2020-06-01 | 383 |
| AY.46 | Nairobi | 2021-02-09 | Denmark 20.0%, United Kingdom 11.0%, Germany 10.0%, Sweden 8.0%, United States of America 7.0% | 2020-06-01 | 253 |
| AY.46 | Nyamira | 2021-09-13 | Denmark 20.0%, United Kingdom 11.0%, Germany 10.0%, Sweden 8.0%, United States of America 7.0% | 2020-06-01 | 469 |
| AY.46 | Nyeri | 2021-07-01 | Denmark 20.0%, United Kingdom 11.0%, Germany 10.0%, Sweden 8.0%, United States of America 7.0% | 2020-06-01 | 395 |
| AY.46 | Siaya | 2021-07-19 | Denmark 20.0%, United Kingdom 11.0%, Germany 10.0%, Sweden 8.0%, United States of America 7.0% | 2020-06-01 | 413 |
| AY.46 | Taita Taveta | 2021-07-13 | Denmark 20.0%, United Kingdom 11.0%, Germany 10.0%, Sweden 8.0%, United States of America 7.0% | 2020-06-01 | 407 |
| AY.46 | Uasin Gishu | 2021-09-21 | Denmark 20.0%, United Kingdom 11.0%, Germany 10.0%, Sweden 8.0%, United States of America 7.0% | 2020-06-01 | 477 |
| AY.46 | Vihiga | 2021-09-30 | Denmark 20.0%, United Kingdom 11.0%, Germany 10.0%, Sweden 8.0%, United States of America 7.0% | 2020-06-01 | 486 |
| AY.46.2 | Busia | 2021-07-29 | Germany 36.0%, Turkey 29.0%, United States of America 5.0%, Sweden 5.0%, France 4.0% | 2021-03-09 | 142 |
| AY.46.2 | Garissa | 2021-08-24 | Germany 36.0%, Turkey 29.0%, United States of America 5.0%, Sweden 5.0%, France 4.0% | 2021-03-09 | 168 |
| AY.46.2 | Kiambu | 2021-07-20 | Germany 36.0%, Turkey 29.0%, United States of America 5.0%, Sweden 5.0%, France 4.0% | 2021-03-09 | 133 |
| AY.46.2 | Kilifi | 2021-07-04 | Germany 36.0%, Turkey 29.0%, United States of America 5.0%, Sweden 5.0%, France 4.0% | 2021-03-09 | 117 |
| AY.46.2 | Laikipia | 2021-07-07 | Germany 36.0%, Turkey 29.0%, United States of America 5.0%, Sweden 5.0%, France 4.0% | 2021-03-09 | 120 |
| AY.46.2 | Lamu | 2021-07-09 | Germany 36.0%, Turkey 29.0%, United States of America 5.0%, Sweden 5.0%, France 4.0% | 2021-03-09 | 122 |
| AY.46.2 | Machakos | 2021-07-14 | Germany 36.0%, Turkey 29.0%, United States of America 5.0%, Sweden 5.0%, France 4.0% | 2021-03-09 | 127 |
| AY.46.2 | Mombasa | 2021-06-23 | Germany 36.0%, Turkey 29.0%, United States of America 5.0%, Sweden 5.0%, France 4.0% | 2021-03-09 | 106 |
| AY.46.2 | Nairobi | 2021-07-08 | Germany 36.0%, Turkey 29.0%, United States of America 5.0%, Sweden 5.0%, France 4.0% | 2021-03-09 | 121 |
| AY.46.2 | Taita Taveta | 2021-07-13 | Germany 36.0%, Turkey 29.0%, United States of America 5.0%, Sweden 5.0%, France 4.0% | 2021-03-09 | 126 |
| AY.46.4 | Baringo | 2021-10-12 | United States of America 93.0%, Denmark 1.0%, Turkey 1.0%, Uganda 1.0%, Belgium 1.0% | 2021-03-29 | 197 |
| AY.46.4 | Kericho | 2021-08-24 | United States of America 93.0%, Denmark 1.0%, Turkey 1.0%, Uganda 1.0%, Belgium 1.0% | 2021-03-29 | 148 |
| AY.46.4 | Kiambu | 2021-08-31 | United States of America 93.0%, Denmark 1.0%, Turkey 1.0%, Uganda 1.0%, Belgium 1.0% | 2021-03-29 | 155 |
| AY.46.4 | Kisii | 2021-09-16 | United States of America 93.0%, Denmark 1.0%, Turkey 1.0%, Uganda 1.0%, Belgium 1.0% | 2021-03-29 | 171 |
| AY.46.4 | Lamu | 2021-10-05 | United States of America 93.0%, Denmark 1.0%, Turkey 1.0%, Uganda 1.0%, Belgium 1.0% | 2021-03-29 | 190 |
| AY.46.4 | Machakos | 2021-06-08 | United States of America 93.0%, Denmark 1.0%, Turkey 1.0%, Uganda 1.0%, Belgium 1.0% | 2021-03-29 | 71 |
| AY.46.4 | Nairobi | 2021-02-09 | United States of America 93.0%, Denmark 1.0%, Turkey 1.0%, Uganda 1.0%, Belgium 1.0% | 2021-03-29 | -48 |
| AY.46.5 | Machakos | 2021-07-29 | United Kingdom 97.0%, Ireland 1.0%, Sweden 0.0%, Spain 0.0%, Germany 0.0% | 2021-02-06 | 173 |
| AY.46.6 | Machakos | 2021-09-15 | Germany 27.0%, Switzerland 18.0%, Italy 8.0%, Sweden 7.0%, Slovenia 7.0% | 2020-08-11 | 400 |
| AY.46.6 | Mombasa | 2021-09-24 | Germany 27.0%, Switzerland 18.0%, Italy 8.0%, Sweden 7.0%, Slovenia 7.0% | 2020-08-11 | 409 |
| AY.46.6 | Nairobi | 2021-02-09 | Germany 27.0%, Switzerland 18.0%, Italy 8.0%, Sweden 7.0%, Slovenia 7.0% | 2020-08-11 | 182 |
| AY.5 | Machakos | 2021-09-17 | United Kingdom 65.0%, France 5.0%, Portugal 4.0%, United States of America 4.0%, Ireland 3.0% | 2020-11-15 | 306 |
| AY.55 | Mombasa | 2021-06-30 | India 55.0%, Rwanda 17.0%, Germany 6.0%, United States of America 4.0%, Spain 3.0% | 2021-01-07 | 174 |
| AY.79 | Kilifi | 2021-08-03 | Malaysia 66.0%, Indonesia 21.0%, Thailand 8.0%, Singapore 2.0%, Vietnam 1.0% | 2021-01-03 | 212 |
| AY.79 | Machakos | 2021-08-31 | Malaysia 66.0%, Indonesia 21.0%, Thailand 8.0%, Singapore 2.0%, Vietnam 1.0% | 2021-01-03 | 240 |
| AY.79 | Nyamira | 2021-09-16 | Malaysia 66.0%, Indonesia 21.0%, Thailand 8.0%, Singapore 2.0%, Vietnam 1.0% | 2021-01-03 | 256 |
| AY.9.2 | Kakamega | 2021-11-03 | Germany 14.0%, Netherlands 14.0%, United Kingdom 8.0%, France 7.0%, Sweden 7.0% | 2020-07-16 | 475 |
| AY.91 | Mombasa | 2021-06-16 | South_Africa 31.0%, Germany 27.0%, Angola 8.0%, Namibia 6.0%, Portugal 6.0% | 2021-02-08 | 128 |
| AY.98 | Nairobi | 2021-04-29 | United Kingdom 91.0%, United States of America 2.0%, Germany 1.0%, Ireland 1.0%, Denmark 1.0% | 2020-10-14 | 197 |
| B.1.1.7 | Baringo | 2021-04-29 | United Kingdom 24.0%, United States of America 20.0%, Germany 9.0%, Sweden 6.0%, Denmark 6.0% | 2020-09-03 | 238 |
| B.1.1.7 | Bungoma | 2021-03-22 | United Kingdom 24.0%, United States of America 20.0%, Germany 9.0%, Sweden 6.0%, Denmark 6.0% | 2020-09-03 | 200 |
| B.1.1.7 | Busia | 2021-04-22 | United Kingdom 24.0%, United States of America 20.0%, Germany 9.0%, Sweden 6.0%, Denmark 6.0% | 2020-09-03 | 231 |
| B.1.1.7 | Embu | 2021-03-16 | United Kingdom 24.0%, United States of America 20.0%, Germany 9.0%, Sweden 6.0%, Denmark 6.0% | 2020-09-03 | 194 |
| B.1.1.7 | Garissa | 2021-03-21 | United Kingdom 24.0%, United States of America 20.0%, Germany 9.0%, Sweden 6.0%, Denmark 6.0% | 2020-09-03 | 199 |
| B.1.1.7 | Homa Bay | 2021-03-22 | United Kingdom 24.0%, United States of America 20.0%, Germany 9.0%, Sweden 6.0%, Denmark 6.0% | 2020-09-03 | 200 |
| B.1.1.7 | Kajiado | 2021-01-12 | United Kingdom 24.0%, United States of America 20.0%, Germany 9.0%, Sweden 6.0%, Denmark 6.0% | 2020-09-03 | 131 |
| B.1.1.7 | Kakamega | 2021-04-26 | United Kingdom 24.0%, United States of America 20.0%, Germany 9.0%, Sweden 6.0%, Denmark 6.0% | 2020-09-03 | 235 |
| B.1.1.7 | Kericho | 2021-06-11 | United Kingdom 24.0%, United States of America 20.0%, Germany 9.0%, Sweden 6.0%, Denmark 6.0% | 2020-09-03 | 281 |
| B.1.1.7 | Kiambu | 2021-01-03 | United Kingdom 24.0%, United States of America 20.0%, Germany 9.0%, Sweden 6.0%, Denmark 6.0% | 2020-09-03 | 122 |
| B.1.1.7 | Kilifi | 2021-03-03 | United Kingdom 24.0%, United States of America 20.0%, Germany 9.0%, Sweden 6.0%, Denmark 6.0% | 2020-09-03 | 181 |
| B.1.1.7 | Kirinyaga | 2021-03-12 | United Kingdom 24.0%, United States of America 20.0%, Germany 9.0%, Sweden 6.0%, Denmark 6.0% | 2020-09-03 | 190 |
| B.1.1.7 | Kisii | 2021-03-08 | United Kingdom 24.0%, United States of America 20.0%, Germany 9.0%, Sweden 6.0%, Denmark 6.0% | 2020-09-03 | 186 |
| B.1.1.7 | Kisumu | 2021-02-02 | United Kingdom 24.0%, United States of America 20.0%, Germany 9.0%, Sweden 6.0%, Denmark 6.0% | 2020-09-03 | 152 |
| B.1.1.7 | Kwale | 2021-03-04 | United Kingdom 24.0%, United States of America 20.0%, Germany 9.0%, Sweden 6.0%, Denmark 6.0% | 2020-09-03 | 182 |
| B.1.1.7 | Laikipia | 2021-02-09 | United Kingdom 24.0%, United States of America 20.0%, Germany 9.0%, Sweden 6.0%, Denmark 6.0% | 2020-09-03 | 159 |
| B.1.1.7 | Lamu | 2021-03-17 | United Kingdom 24.0%, United States of America 20.0%, Germany 9.0%, Sweden 6.0%, Denmark 6.0% | 2020-09-03 | 195 |
| B.1.1.7 | Machakos | 2021-05-03 | United Kingdom 24.0%, United States of America 20.0%, Germany 9.0%, Sweden 6.0%, Denmark 6.0% | 2020-09-03 | 242 |
| B.1.1.7 | Mandera | 2021-05-07 | United Kingdom 24.0%, United States of America 20.0%, Germany 9.0%, Sweden 6.0%, Denmark 6.0% | 2020-09-03 | 246 |
| B.1.1.7 | Meru | 2021-03-19 | United Kingdom 24.0%, United States of America 20.0%, Germany 9.0%, Sweden 6.0%, Denmark 6.0% | 2020-09-03 | 197 |
| B.1.1.7 | Migori | 2021-03-17 | United Kingdom 24.0%, United States of America 20.0%, Germany 9.0%, Sweden 6.0%, Denmark 6.0% | 2020-09-03 | 195 |
| B.1.1.7 | Mombasa | 2021-01-11 | United Kingdom 24.0%, United States of America 20.0%, Germany 9.0%, Sweden 6.0%, Denmark 6.0% | 2020-09-03 | 130 |
| B.1.1.7 | Murang'a | 2021-02-28 | United Kingdom 24.0%, United States of America 20.0%, Germany 9.0%, Sweden 6.0%, Denmark 6.0% | 2020-09-03 | 178 |
| B.1.1.7 | Nairobi | 2020-09-09 | United Kingdom 24.0%, United States of America 20.0%, Germany 9.0%, Sweden 6.0%, Denmark 6.0% | 2020-09-03 | 6 |
| B.1.1.7 | Nakuru | 2021-03-08 | United Kingdom 24.0%, United States of America 20.0%, Germany 9.0%, Sweden 6.0%, Denmark 6.0% | 2020-09-03 | 186 |
| B.1.1.7 | Nyamira | 2021-03-22 | United Kingdom 24.0%, United States of America 20.0%, Germany 9.0%, Sweden 6.0%, Denmark 6.0% | 2020-09-03 | 200 |
| B.1.1.7 | Nyandarua | 2021-03-16 | United Kingdom 24.0%, United States of America 20.0%, Germany 9.0%, Sweden 6.0%, Denmark 6.0% | 2020-09-03 | 194 |
| B.1.1.7 | Nyeri | 2021-05-25 | United Kingdom 24.0%, United States of America 20.0%, Germany 9.0%, Sweden 6.0%, Denmark 6.0% | 2020-09-03 | 264 |
| B.1.1.7 | Siaya | 2021-03-13 | United Kingdom 24.0%, United States of America 20.0%, Germany 9.0%, Sweden 6.0%, Denmark 6.0% | 2020-09-03 | 191 |
| B.1.1.7 | Taita Taveta | 2021-02-10 | United Kingdom 24.0%, United States of America 20.0%, Germany 9.0%, Sweden 6.0%, Denmark 6.0% | 2020-09-03 | 160 |
| B.1.1.7 | Tana River | 2021-04-15 | United Kingdom 24.0%, United States of America 20.0%, Germany 9.0%, Sweden 6.0%, Denmark 6.0% | 2020-09-03 | 224 |
| B.1.1.7 | Trans Nzoia | 2021-03-11 | United Kingdom 24.0%, United States of America 20.0%, Germany 9.0%, Sweden 6.0%, Denmark 6.0% | 2020-09-03 | 189 |
| B.1.1.7 | Vihiga | 2021-03-08 | United Kingdom 24.0%, United States of America 20.0%, Germany 9.0%, Sweden 6.0%, Denmark 6.0% | 2020-09-03 | 186 |
| B.1.351 | Bomet | 2021-07-30 | South_Africa 19.0%, Philippines 9.0%, United States of America 9.0%, Sweden 8.0%, Germany 7.0% | 2020-09-01 | 332 |
| B.1.351 | Bungoma | 2021-03-25 | South_Africa 19.0%, Philippines 9.0%, United States of America 9.0%, Sweden 8.0%, Germany 7.0% | 2020-09-01 | 205 |
| B.1.351 | Kiambu | 2021-02-12 | South_Africa 19.0%, Philippines 9.0%, United States of America 9.0%, Sweden 8.0%, Germany 7.0% | 2020-09-01 | 164 |
| B.1.351 | Kilifi | 2020-12-15 | South_Africa 19.0%, Philippines 9.0%, United States of America 9.0%, Sweden 8.0%, Germany 7.0% | 2020-09-01 | 105 |
| B.1.351 | Kirinyaga | 2021-03-19 | South_Africa 19.0%, Philippines 9.0%, United States of America 9.0%, Sweden 8.0%, Germany 7.0% | 2020-09-01 | 199 |
| B.1.351 | Kisumu | 2021-03-08 | South_Africa 19.0%, Philippines 9.0%, United States of America 9.0%, Sweden 8.0%, Germany 7.0% | 2020-09-01 | 188 |
| B.1.351 | Kwale | 2021-01-16 | South_Africa 19.0%, Philippines 9.0%, United States of America 9.0%, Sweden 8.0%, Germany 7.0% | 2020-09-01 | 137 |
| B.1.351 | Lamu | 2021-02-16 | South_Africa 19.0%, Philippines 9.0%, United States of America 9.0%, Sweden 8.0%, Germany 7.0% | 2020-09-01 | 168 |
| B.1.351 | Machakos | 2021-06-16 | South_Africa 19.0%, Philippines 9.0%, United States of America 9.0%, Sweden 8.0%, Germany 7.0% | 2020-09-01 | 288 |
| B.1.351 | Migori | 2021-02-24 | South_Africa 19.0%, Philippines 9.0%, United States of America 9.0%, Sweden 8.0%, Germany 7.0% | 2020-09-01 | 176 |
| B.1.351 | Mombasa | 2021-02-13 | South_Africa 19.0%, Philippines 9.0%, United States of America 9.0%, Sweden 8.0%, Germany 7.0% | 2020-09-01 | 165 |
| B.1.351 | Murang'a | 2021-02-09 | South_Africa 19.0%, Philippines 9.0%, United States of America 9.0%, Sweden 8.0%, Germany 7.0% | 2020-09-01 | 161 |
| B.1.351 | Nairobi | 2021-02-04 | South_Africa 19.0%, Philippines 9.0%, United States of America 9.0%, Sweden 8.0%, Germany 7.0% | 2020-09-01 | 156 |
| B.1.351 | Nyandarua | 2021-03-19 | South_Africa 19.0%, Philippines 9.0%, United States of America 9.0%, Sweden 8.0%, Germany 7.0% | 2020-09-01 | 199 |
| B.1.351 | Siaya | 2021-04-18 | South_Africa 19.0%, Philippines 9.0%, United States of America 9.0%, Sweden 8.0%, Germany 7.0% | 2020-09-01 | 229 |
| B.1.351 | Taita Taveta | 2021-01-20 | South_Africa 19.0%, Philippines 9.0%, United States of America 9.0%, Sweden 8.0%, Germany 7.0% | 2020-09-01 | 141 |
| B.1.617.2 | Bomet | 2021-07-27 | United States of America 21.0%, India 18.0%, United Kingdom 13.0%, Turkey 10.0%, Germany 6.0% | 2021-03-01 | 148 |
| B.1.617.2 | Busia | 2021-07-30 | United States of America 21.0%, India 18.0%, United Kingdom 13.0%, Turkey 10.0%, Germany 6.0% | 2021-03-01 | 151 |
| B.1.617.2 | Garissa | 2021-08-26 | United States of America 21.0%, India 18.0%, United Kingdom 13.0%, Turkey 10.0%, Germany 6.0% | 2021-03-01 | 178 |
| B.1.617.2 | Kericho | 2021-08-03 | United States of America 21.0%, India 18.0%, United Kingdom 13.0%, Turkey 10.0%, Germany 6.0% | 2021-03-01 | 155 |
| B.1.617.2 | Kiambu | 2021-07-06 | United States of America 21.0%, India 18.0%, United Kingdom 13.0%, Turkey 10.0%, Germany 6.0% | 2021-03-01 | 127 |
| B.1.617.2 | Kilifi | 2021-05-04 | United States of America 21.0%, India 18.0%, United Kingdom 13.0%, Turkey 10.0%, Germany 6.0% | 2021-03-01 | 64 |
| B.1.617.2 | Kisii | 2021-09-27 | United States of America 21.0%, India 18.0%, United Kingdom 13.0%, Turkey 10.0%, Germany 6.0% | 2021-03-01 | 210 |
| B.1.617.2 | Kisumu | 2021-05-21 | United States of America 21.0%, India 18.0%, United Kingdom 13.0%, Turkey 10.0%, Germany 6.0% | 2021-03-01 | 81 |
| B.1.617.2 | Laikipia | 2021-06-25 | United States of America 21.0%, India 18.0%, United Kingdom 13.0%, Turkey 10.0%, Germany 6.0% | 2021-03-01 | 116 |
| B.1.617.2 | Lamu | 2021-06-30 | United States of America 21.0%, India 18.0%, United Kingdom 13.0%, Turkey 10.0%, Germany 6.0% | 2021-03-01 | 121 |
| B.1.617.2 | Machakos | 2021-05-18 | United States of America 21.0%, India 18.0%, United Kingdom 13.0%, Turkey 10.0%, Germany 6.0% | 2021-03-01 | 78 |
| B.1.617.2 | Migori | 2021-06-05 | United States of America 21.0%, India 18.0%, United Kingdom 13.0%, Turkey 10.0%, Germany 6.0% | 2021-03-01 | 96 |
| B.1.617.2 | Mombasa | 2021-05-03 | United States of America 21.0%, India 18.0%, United Kingdom 13.0%, Turkey 10.0%, Germany 6.0% | 2021-03-01 | 63 |
| B.1.617.2 | Nairobi | 2021-04-09 | United States of America 21.0%, India 18.0%, United Kingdom 13.0%, Turkey 10.0%, Germany 6.0% | 2021-03-01 | 39 |
| B.1.617.2 | Nyamira | 2021-09-24 | United States of America 21.0%, India 18.0%, United Kingdom 13.0%, Turkey 10.0%, Germany 6.0% | 2021-03-01 | 207 |
| B.1.617.2 | Siaya | 2021-07-04 | United States of America 21.0%, India 18.0%, United Kingdom 13.0%, Turkey 10.0%, Germany 6.0% | 2021-03-01 | 125 |
| B.1.617.2 | Taita Taveta | 2021-06-15 | United States of America 21.0%, India 18.0%, United Kingdom 13.0%, Turkey 10.0%, Germany 6.0% | 2021-03-01 | 106 |
| BA.1 | Kajiado | 2021-12-04 | United Kingdom 41.0%, United States of America 22.0%, Denmark 5.0%, Germany 4.0%, Brazil 3.0% | 2021-09-02 | 93 |
| BA.1 | Kakamega | 2021-12-25 | United Kingdom 41.0%, United States of America 22.0%, Denmark 5.0%, Germany 4.0%, Brazil 3.0% | 2021-09-02 | 114 |
| BA.1 | Kiambu | 2021-12-11 | United Kingdom 41.0%, United States of America 22.0%, Denmark 5.0%, Germany 4.0%, Brazil 3.0% | 2021-09-02 | 100 |
| BA.1 | Kilifi | 2021-12-15 | United Kingdom 41.0%, United States of America 22.0%, Denmark 5.0%, Germany 4.0%, Brazil 3.0% | 2021-09-02 | 104 |
| BA.1 | Kisumu | 2021-12-16 | United Kingdom 41.0%, United States of America 22.0%, Denmark 5.0%, Germany 4.0%, Brazil 3.0% | 2021-09-02 | 105 |
| BA.1 | Kwale | 2021-12-20 | United Kingdom 41.0%, United States of America 22.0%, Denmark 5.0%, Germany 4.0%, Brazil 3.0% | 2021-09-02 | 109 |
| BA.1 | Machakos | 2021-12-15 | United Kingdom 41.0%, United States of America 22.0%, Denmark 5.0%, Germany 4.0%, Brazil 3.0% | 2021-09-02 | 104 |
| BA.1 | Migori | 2021-12-22 | United Kingdom 41.0%, United States of America 22.0%, Denmark 5.0%, Germany 4.0%, Brazil 3.0% | 2021-09-02 | 111 |
| BA.1 | Mombasa | 2021-12-14 | United Kingdom 41.0%, United States of America 22.0%, Denmark 5.0%, Germany 4.0%, Brazil 3.0% | 2021-09-02 | 103 |
| BA.1 | Nairobi | 2021-11-27 | United Kingdom 41.0%, United States of America 22.0%, Denmark 5.0%, Germany 4.0%, Brazil 3.0% | 2021-09-02 | 86 |
| BA.1 | Nakuru | 2021-12-09 | United Kingdom 41.0%, United States of America 22.0%, Denmark 5.0%, Germany 4.0%, Brazil 3.0% | 2021-09-02 | 98 |
| BA.1 | Siaya | 2021-12-04 | United Kingdom 41.0%, United States of America 22.0%, Denmark 5.0%, Germany 4.0%, Brazil 3.0% | 2021-09-02 | 93 |
| BA.1 | Uasin Gishu | 2022-01-08 | United Kingdom 41.0%, United States of America 22.0%, Denmark 5.0%, Germany 4.0%, Brazil 3.0% | 2021-09-02 | 128 |
| BA.1.1 | Bungoma | 2021-12-14 | United States of America 48.0%, United Kingdom 21.0%, Germany 7.0%, Canada 4.0%, France 2.0% | 2021-09-09 | 96 |
| BA.1.1 | Busia | 2021-12-09 | United States of America 48.0%, United Kingdom 21.0%, Germany 7.0%, Canada 4.0%, France 2.0% | 2021-09-09 | 91 |
| BA.1.1 | Garissa | 2021-12-15 | United States of America 48.0%, United Kingdom 21.0%, Germany 7.0%, Canada 4.0%, France 2.0% | 2021-09-09 | 97 |
| BA.1.1 | Homa Bay | 2021-12-17 | United States of America 48.0%, United Kingdom 21.0%, Germany 7.0%, Canada 4.0%, France 2.0% | 2021-09-09 | 99 |
| BA.1.1 | Kakamega | 2021-12-10 | United States of America 48.0%, United Kingdom 21.0%, Germany 7.0%, Canada 4.0%, France 2.0% | 2021-09-09 | 92 |
| BA.1.1 | Kericho | 2021-12-15 | United States of America 48.0%, United Kingdom 21.0%, Germany 7.0%, Canada 4.0%, France 2.0% | 2021-09-09 | 97 |
| BA.1.1 | Kiambu | 2021-12-11 | United States of America 48.0%, United Kingdom 21.0%, Germany 7.0%, Canada 4.0%, France 2.0% | 2021-09-09 | 93 |
| BA.1.1 | Kilifi | 2021-12-15 | United States of America 48.0%, United Kingdom 21.0%, Germany 7.0%, Canada 4.0%, France 2.0% | 2021-09-09 | 97 |
| BA.1.1 | Kisii | 2021-12-13 | United States of America 48.0%, United Kingdom 21.0%, Germany 7.0%, Canada 4.0%, France 2.0% | 2021-09-09 | 95 |
| BA.1.1 | Kisumu | 2021-12-14 | United States of America 48.0%, United Kingdom 21.0%, Germany 7.0%, Canada 4.0%, France 2.0% | 2021-09-09 | 96 |
| BA.1.1 | Kwale | 2021-12-20 | United States of America 48.0%, United Kingdom 21.0%, Germany 7.0%, Canada 4.0%, France 2.0% | 2021-09-09 | 102 |
| BA.1.1 | Laikipia | 2021-12-01 | United States of America 48.0%, United Kingdom 21.0%, Germany 7.0%, Canada 4.0%, France 2.0% | 2021-09-09 | 83 |
| BA.1.1 | Lamu | 2021-12-27 | United States of America 48.0%, United Kingdom 21.0%, Germany 7.0%, Canada 4.0%, France 2.0% | 2021-09-09 | 109 |
| BA.1.1 | Machakos | 2021-12-10 | United States of America 48.0%, United Kingdom 21.0%, Germany 7.0%, Canada 4.0%, France 2.0% | 2021-09-09 | 92 |
| BA.1.1 | Migori | 2021-12-17 | United States of America 48.0%, United Kingdom 21.0%, Germany 7.0%, Canada 4.0%, France 2.0% | 2021-09-09 | 99 |
| BA.1.1 | Mombasa | 2021-12-09 | United States of America 48.0%, United Kingdom 21.0%, Germany 7.0%, Canada 4.0%, France 2.0% | 2021-09-09 | 91 |
| BA.1.1 | Murang'a | 2021-12-20 | United States of America 48.0%, United Kingdom 21.0%, Germany 7.0%, Canada 4.0%, France 2.0% | 2021-09-09 | 102 |
| BA.1.1 | Nairobi | 2021-11-14 | United States of America 48.0%, United Kingdom 21.0%, Germany 7.0%, Canada 4.0%, France 2.0% | 2021-09-09 | 66 |
| BA.1.1 | Nakuru | 2021-12-20 | United States of America 48.0%, United Kingdom 21.0%, Germany 7.0%, Canada 4.0%, France 2.0% | 2021-09-09 | 102 |
| BA.1.1 | Narok | 2021-12-18 | United States of America 48.0%, United Kingdom 21.0%, Germany 7.0%, Canada 4.0%, France 2.0% | 2021-09-09 | 100 |
| BA.1.1 | Nyamira | 2022-01-19 | United States of America 48.0%, United Kingdom 21.0%, Germany 7.0%, Canada 4.0%, France 2.0% | 2021-09-09 | 132 |
| BA.1.1 | Nyeri | 2021-12-29 | United States of America 48.0%, United Kingdom 21.0%, Germany 7.0%, Canada 4.0%, France 2.0% | 2021-09-09 | 111 |
| BA.1.1 | Siaya | 2022-01-10 | United States of America 48.0%, United Kingdom 21.0%, Germany 7.0%, Canada 4.0%, France 2.0% | 2021-09-09 | 123 |
| BA.1.1 | Taita Taveta | 2021-12-20 | United States of America 48.0%, United Kingdom 21.0%, Germany 7.0%, Canada 4.0%, France 2.0% | 2021-09-09 | 102 |
| BA.1.1 | Trans Nzoia | 2021-12-17 | United States of America 48.0%, United Kingdom 21.0%, Germany 7.0%, Canada 4.0%, France 2.0% | 2021-09-09 | 99 |
| BA.1.1 | Uasin Gishu | 2022-01-04 | United States of America 48.0%, United Kingdom 21.0%, Germany 7.0%, Canada 4.0%, France 2.0% | 2021-09-09 | 117 |
| BA.1.1 | Vihiga | 2022-01-03 | United States of America 48.0%, United Kingdom 21.0%, Germany 7.0%, Canada 4.0%, France 2.0% | 2021-09-09 | 116 |
| BA.1.1.1 | Bungoma | 2021-12-14 | France 25.0%, Germany 18.0%, United Kingdom 11.0%, Japan 10.0%, Spain 6.0% | 2021-11-12 | 32 |
| BA.1.1.1 | Garissa | 2022-01-23 | France 25.0%, Germany 18.0%, United Kingdom 11.0%, Japan 10.0%, Spain 6.0% | 2021-11-12 | 72 |
| BA.1.1.1 | Kakamega | 2021-12-18 | France 25.0%, Germany 18.0%, United Kingdom 11.0%, Japan 10.0%, Spain 6.0% | 2021-11-12 | 36 |
| BA.1.1.1 | Kiambu | 2021-12-14 | France 25.0%, Germany 18.0%, United Kingdom 11.0%, Japan 10.0%, Spain 6.0% | 2021-11-12 | 32 |
| BA.1.1.1 | Kilifi | 2021-12-15 | France 25.0%, Germany 18.0%, United Kingdom 11.0%, Japan 10.0%, Spain 6.0% | 2021-11-12 | 33 |
| BA.1.1.1 | Kisumu | 2021-12-17 | France 25.0%, Germany 18.0%, United Kingdom 11.0%, Japan 10.0%, Spain 6.0% | 2021-11-12 | 35 |
| BA.1.1.1 | Kwale | 2021-12-21 | France 25.0%, Germany 18.0%, United Kingdom 11.0%, Japan 10.0%, Spain 6.0% | 2021-11-12 | 39 |
| BA.1.1.1 | Machakos | 2021-12-14 | France 25.0%, Germany 18.0%, United Kingdom 11.0%, Japan 10.0%, Spain 6.0% | 2021-11-12 | 32 |
| BA.1.1.1 | Migori | 2021-12-21 | France 25.0%, Germany 18.0%, United Kingdom 11.0%, Japan 10.0%, Spain 6.0% | 2021-11-12 | 39 |
| BA.1.1.1 | Mombasa | 2021-12-24 | France 25.0%, Germany 18.0%, United Kingdom 11.0%, Japan 10.0%, Spain 6.0% | 2021-11-12 | 42 |
| BA.1.1.1 | Nairobi | 2021-12-02 | France 25.0%, Germany 18.0%, United Kingdom 11.0%, Japan 10.0%, Spain 6.0% | 2021-11-12 | 20 |
| BA.1.1.1 | Uasin Gishu | 2022-01-05 | France 25.0%, Germany 18.0%, United Kingdom 11.0%, Japan 10.0%, Spain 6.0% | 2021-11-12 | 54 |
| BA.1.1.11 | Nairobi | 2021-12-31 | Switzerland 53.0%, Germany 14.0%, Italy 9.0%, United Kingdom 6.0%, France 3.0% | 2021-11-30 | 31 |
| BA.1.1.14 | Kwale | 2021-12-26 | United Kingdom 54.0%, United States of America 16.0%, Germany 6.0%, Spain 5.0%, Brazil 4.0% | 2021-11-25 | 31 |
| BA.1.1.14 | Mombasa | 2021-12-24 | United Kingdom 54.0%, United States of America 16.0%, Germany 6.0%, Spain 5.0%, Brazil 4.0% | 2021-11-25 | 29 |
| BA.1.1.14 | Nairobi | 2022-01-23 | United Kingdom 54.0%, United States of America 16.0%, Germany 6.0%, Spain 5.0%, Brazil 4.0% | 2021-11-25 | 59 |
| BA.1.1.4 | Bungoma | 2021-12-21 | United Kingdom 83.0%, Sweden 5.0%, Norway 3.0%, Slovakia 2.0%, United States of America 2.0% | 2021-12-13 | 8 |
| BA.1.1.4 | Kisumu | 2021-12-29 | United Kingdom 83.0%, Sweden 5.0%, Norway 3.0%, Slovakia 2.0%, United States of America 2.0% | 2021-12-13 | 16 |
| BA.1.1.4 | Vihiga | 2021-12-22 | United Kingdom 83.0%, Sweden 5.0%, Norway 3.0%, Slovakia 2.0%, United States of America 2.0% | 2021-12-13 | 9 |
| BA.1.10 | Nairobi | 2021-12-15 | United Kingdom 74.0%, Croatia 9.0%, Slovenia 4.0%, Germany 2.0%, South_Africa 2.0% | 2021-11-24 | 21 |
| BA.1.13 | Nairobi | 2021-11-29 | France 28.0%, Sweden 12.0%, Germany 12.0%, United States of America 7.0%, United Kingdom 6.0% | 2021-11-16 | 13 |
| BA.1.14 | Kakamega | 2021-12-16 | Belgium 21.0%, Brazil 19.0%, United Kingdom 12.0%, Denmark 10.0%, Germany 7.0% | 2021-10-20 | 57 |
| BA.1.14 | Kiambu | 2021-12-11 | Belgium 21.0%, Brazil 19.0%, United Kingdom 12.0%, Denmark 10.0%, Germany 7.0% | 2021-10-20 | 52 |
| BA.1.14 | Migori | 2021-12-25 | Belgium 21.0%, Brazil 19.0%, United Kingdom 12.0%, Denmark 10.0%, Germany 7.0% | 2021-10-20 | 66 |
| BA.1.14 | Mombasa | 2021-12-24 | Belgium 21.0%, Brazil 19.0%, United Kingdom 12.0%, Denmark 10.0%, Germany 7.0% | 2021-10-20 | 65 |
| BA.1.14 | Nairobi | 2021-12-07 | Belgium 21.0%, Brazil 19.0%, United Kingdom 12.0%, Denmark 10.0%, Germany 7.0% | 2021-10-20 | 48 |
| BA.1.14 | Uasin Gishu | 2022-01-07 | Belgium 21.0%, Brazil 19.0%, United Kingdom 12.0%, Denmark 10.0%, Germany 7.0% | 2021-10-20 | 79 |
| BA.1.15 | Kisumu | 2021-12-16 | United States of America 69.0%, United Kingdom 14.0%, Canada 4.0%, Germany 2.0%, Japan 1.0% | 2021-09-12 | 95 |
| BA.1.15 | Kwale | 2021-12-22 | United States of America 69.0%, United Kingdom 14.0%, Canada 4.0%, Germany 2.0%, Japan 1.0% | 2021-09-12 | 101 |
| BA.1.15 | Machakos | 2021-12-20 | United States of America 69.0%, United Kingdom 14.0%, Canada 4.0%, Germany 2.0%, Japan 1.0% | 2021-09-12 | 99 |
| BA.1.15 | Mombasa | 2021-12-13 | United States of America 69.0%, United Kingdom 14.0%, Canada 4.0%, Germany 2.0%, Japan 1.0% | 2021-09-12 | 92 |
| BA.1.15 | Nairobi | 2021-12-12 | United States of America 69.0%, United Kingdom 14.0%, Canada 4.0%, Germany 2.0%, Japan 1.0% | 2021-09-12 | 91 |
| BA.1.15.1 | Nairobi | 2021-12-11 | United Kingdom 87.0%, Germany 3.0%, United States of America 2.0%, France 1.0%, Poland 1.0% | 2021-11-18 | 23 |
| BA.1.17 | Kwale | 2021-12-22 | Australia 27.0%, United Kingdom 16.0%, Spain 15.0%, United States of America 8.0%, Germany 7.0% | 2021-09-28 | 85 |
| BA.1.17 | Laikipia | 2021-12-19 | Australia 27.0%, United Kingdom 16.0%, Spain 15.0%, United States of America 8.0%, Germany 7.0% | 2021-09-28 | 82 |
| BA.1.17 | Mombasa | 2021-12-11 | Australia 27.0%, United Kingdom 16.0%, Spain 15.0%, United States of America 8.0%, Germany 7.0% | 2021-09-28 | 74 |
| BA.1.17 | Nairobi | 2021-11-28 | Australia 27.0%, United Kingdom 16.0%, Spain 15.0%, United States of America 8.0%, Germany 7.0% | 2021-09-28 | 61 |
| BA.1.17 | Uasin Gishu | 2022-01-08 | Australia 27.0%, United Kingdom 16.0%, Spain 15.0%, United States of America 8.0%, Germany 7.0% | 2021-09-28 | 102 |
| BA.1.17.2 | Bungoma | 2021-12-21 | United Kingdom 70.0%, United States of America 6.0%, Canada 4.0%, Germany 3.0%, Denmark 3.0% | 2021-10-01 | 81 |
| BA.1.17.2 | Kisii | 2021-12-21 | United Kingdom 70.0%, United States of America 6.0%, Canada 4.0%, Germany 3.0%, Denmark 3.0% | 2021-10-01 | 81 |
| BA.1.17.2 | Kisumu | 2021-12-28 | United Kingdom 70.0%, United States of America 6.0%, Canada 4.0%, Germany 3.0%, Denmark 3.0% | 2021-10-01 | 88 |
| BA.1.17.2 | Migori | 2021-12-22 | United Kingdom 70.0%, United States of America 6.0%, Canada 4.0%, Germany 3.0%, Denmark 3.0% | 2021-10-01 | 82 |
| BA.1.17.2 | Mombasa | 2021-12-11 | United Kingdom 70.0%, United States of America 6.0%, Canada 4.0%, Germany 3.0%, Denmark 3.0% | 2021-10-01 | 71 |
| BA.1.17.2 | Nairobi | 2021-12-11 | United Kingdom 70.0%, United States of America 6.0%, Canada 4.0%, Germany 3.0%, Denmark 3.0% | 2021-10-01 | 71 |
| BA.1.17.2 | Taita Taveta | 2021-12-21 | United Kingdom 70.0%, United States of America 6.0%, Canada 4.0%, Germany 3.0%, Denmark 3.0% | 2021-10-01 | 81 |
| BA.1.18 | Busia | 2021-12-28 | United States of America 21.0%, France 21.0%, Germany 19.0%, United Kingdom 8.0%, Switzerland 5.0% | 2021-11-02 | 56 |
| BA.1.18 | Garissa | 2021-12-24 | United States of America 21.0%, France 21.0%, Germany 19.0%, United Kingdom 8.0%, Switzerland 5.0% | 2021-11-02 | 52 |
| BA.1.18 | Kilifi | 2021-12-21 | United States of America 21.0%, France 21.0%, Germany 19.0%, United Kingdom 8.0%, Switzerland 5.0% | 2021-11-02 | 49 |
| BA.1.18 | Kirinyaga | 2021-11-30 | United States of America 21.0%, France 21.0%, Germany 19.0%, United Kingdom 8.0%, Switzerland 5.0% | 2021-11-02 | 28 |
| BA.1.18 | Kwale | 2021-12-22 | United States of America 21.0%, France 21.0%, Germany 19.0%, United Kingdom 8.0%, Switzerland 5.0% | 2021-11-02 | 50 |
| BA.1.18 | Mombasa | 2021-12-24 | United States of America 21.0%, France 21.0%, Germany 19.0%, United Kingdom 8.0%, Switzerland 5.0% | 2021-11-02 | 52 |
| BA.1.18 | Nairobi | 2021-11-27 | United States of America 21.0%, France 21.0%, Germany 19.0%, United Kingdom 8.0%, Switzerland 5.0% | 2021-11-02 | 25 |
| BA.1.19 | Bungoma | 2021-12-14 | Denmark 24.0%, United States of America 20.0%, Germany 13.0%, Portugal 11.0%, United Kingdom 9.0% | 2021-11-12 | 32 |
| BA.1.19 | Kilifi | 2021-12-21 | Denmark 24.0%, United States of America 20.0%, Germany 13.0%, Portugal 11.0%, United Kingdom 9.0% | 2021-11-12 | 39 |
| BA.1.19 | Mombasa | 2021-12-06 | Denmark 24.0%, United States of America 20.0%, Germany 13.0%, Portugal 11.0%, United Kingdom 9.0% | 2021-11-12 | 24 |
| BA.1.19 | Nairobi | 2021-12-02 | Denmark 24.0%, United States of America 20.0%, Germany 13.0%, Portugal 11.0%, United Kingdom 9.0% | 2021-11-12 | 20 |
| BA.1.20 | Nairobi | 2022-01-13 | United States of America 83.0%, Canada 6.0%, Germany 2.0%, United Kingdom 1.0%, Poland 1.0% | 2021-09-15 | 120 |
| BA.1.21 | Nairobi | 2021-11-30 | Norway 51.0%, Germany 9.0%, Denmark 5.0%, South_Africa 5.0%, Italy 5.0% | 2021-10-24 | 37 |
| BA.1.5 | Nairobi | 2021-12-12 | United Kingdom 97.0%, Poland 1.0%, United States of America 1.0%, Portugal 1.0%, Brazil 1.0% | 2021-12-12 | 0 |
| BA.1.9 | Kilifi | 2021-12-09 | Brazil 93.0%, United Kingdom 3.0%, United States of America 1.0%, Japan 0.0%, Germany 0.0% | 2021-12-11 | -2 |
| BA.1.9 | Kwale | 2021-12-16 | Brazil 93.0%, United Kingdom 3.0%, United States of America 1.0%, Japan 0.0%, Germany 0.0% | 2021-12-11 | 5 |
| BA.1.9 | Mombasa | 2021-12-14 | Brazil 93.0%, United Kingdom 3.0%, United States of America 1.0%, Japan 0.0%, Germany 0.0% | 2021-12-11 | 3 |
| BA.1.9 | Nairobi | 2021-11-30 | Brazil 93.0%, United Kingdom 3.0%, United States of America 1.0%, Japan 0.0%, Germany 0.0% | 2021-12-11 | -11 |
| BA.1.9 | Taita Taveta | 2021-12-21 | Brazil 93.0%, United Kingdom 3.0%, United States of America 1.0%, Japan 0.0%, Germany 0.0% | 2021-12-11 | 10 |
| BA.2 | Kilifi | 2022-04-20 | United Kingdom 33.0%, Denmark 13.0%, Germany 13.0%, United States of America 11.0%, France 5.0% | 2021-10-11 | 191 |
| BA.2 | Nairobi | 2021-12-14 | United Kingdom 33.0%, Denmark 13.0%, Germany 13.0%, United States of America 11.0%, France 5.0% | 2021-10-11 | 64 |
| BA.2.10 | Nairobi | 2022-05-30 | India 31.0%, United Kingdom 30.0%, United States of America 11.0%, Japan 7.0%, New_Zealand 3.0% | 2021-11-25 | 186 |
| BA.2.12.1 | Nairobi | 2022-06-06 | United States of America 85.0%, Canada 6.0%, United Kingdom 2.0%, Denmark 1.0%, Israel 1.0% | 2021-12-14 | 174 |
| BA.2.3 | Mombasa | 2021-12-24 | United States of America 24.0%, United Kingdom 17.0%, Canada 11.0%, Japan 10.0%, South_Korea 7.0% | 2021-09-29 | 86 |
| BA.2.3 | Nairobi | 2022-01-22 | United States of America 24.0%, United Kingdom 17.0%, Canada 11.0%, Japan 10.0%, South_Korea 7.0% | 2021-09-29 | 115 |
| BA.2.3.2 | Nairobi | 2022-02-24 | Vietnam 30.0%, Japan 28.0%, United States of America 10.0%, Australia 5.0%, United Kingdom 4.0% | 2022-01-23 | 32 |
| BA.2.31 | Kilifi | 2022-05-16 | Israel 60.0%, United States of America 29.0%, United Kingdom 5.0%, Germany 2.0%, Canada 1.0% | 2022-01-12 | 124 |
| BA.2.9 | Nairobi | 2022-03-26 | Denmark 28.0%, Germany 19.0%, United Kingdom 14.0%, United States of America 9.0%, Sweden 4.0% | 2021-12-11 | 105 |
| BA.4 | Kilifi | 2022-06-10 | United States of America 29.0%, United Kingdom 15.0%, South_Africa 15.0%, Israel 8.0%, Denmark 6.0% | 2022-01-10 | 151 |
| BA.4 | Nairobi | 2022-06-03 | United States of America 29.0%, United Kingdom 15.0%, South_Africa 15.0%, Israel 8.0%, Denmark 6.0% | 2022-01-10 | 144 |
| BA.5 | Kilifi | 2022-05-31 | United States of America 26.0%, Germany 16.0%, Israel 12.0%, United Kingdom 11.0%, Denmark 9.0% | 2022-01-05 | 146 |
